# Chromosome 8 gain is associated with high-grade transformation in MPNST

**DOI:** 10.1101/2020.12.04.20242818

**Authors:** Carina Dehner, Chang In Moon, Zhaohe Zhou, Xiyuan Zhang, Chris Miller, Hua Xu, Xiaodan Wan, Kuangying Yang, Jay Mashl, Sara J.C. Gosline, Yuxi Wang, Xiaochun Zhang, Abigail Godec, Paul A. Jones, Sonika Dahiya, Himanshi Bhatia, Tina Primeau, Shunqiang Li, Kai Pollard, Li Ding, Christine A. Pratilas, Jack F. Shern, Angela C. Hirbe

## Abstract

One of the most common malignancies affecting adults with Neurofibromatosis type 1 (NF1) is the malignant peripheral nerve sheath tumor (MPNST), an aggressive and often fatal sarcoma which commonly arises from benign plexiform neurofibromas. Despite advances in our understanding of MPNST pathobiology, there are few effective therapeutic options, and no investigational agents have proven success in clinical trials. To further understand the genomic heterogeneity of MPNST, and to generate a preclinical platform that encompasses this heterogeneity, we developed a collection of NF1-MPNST patient derived xenografts (PDX). These PDX were compared to the primary tumors from which they were derived using copy number analysis, whole exome and RNA sequencing. We identified chromosome 8 gain as a recurrent genomic event in MPNST and validated its occurrence by FISH in the PDX and parental tumors, in a validation cohort, and by single cell sequencing in the PDX. Finally, we show that chromosome 8 gain is associated with inferior overall survival in soft tissue sarcomas. Taken together, these data suggest that chromosome 8 gain is a critical event in MPNST pathogenesis, and may account for the aggressive nature and poor outcomes in this sarcoma subtype.

## Introduction

Malignant peripheral nerve sheath tumors (MPNST) are aggressive soft tissue sarcomas with a devastating prognosis. About half of MPNST arise in patients with an underlying autosomal dominant genetic disorder, Neurofibromatosis type 1 (NF1), while the other half arise de novo or in the setting of prior radiation therapy(1-3). NF1 is one of the most common genetic syndromes with an annual incidence of 1 in 2500 individuals world-wide and the risk of MPNST in a genetically pre-disposed individual is about 8-13% (4). In the context of NF1, MPNST are most often the result of malignant transformation of benign precursor lesions, plexiform neurofibroma (PN) or atypical neurofibroma (AN) (5).

Critical inactivating mutations in the *NF1* gene and consequent hyperactive RAS signaling, are seen in sporadic as well as NF1-associated MPNST (6, 7). Mutations in other genes such as *TP53, EED, SUZ12*, and *CDKN2A* are required for malignant transformation to MPNST (8). Despite advances in understanding the oncogenic drivers of MPNST, chemotherapy is inconsistently effective, and other effective therapies are lacking. The only curative option is aggressive local control with surgery and radiation therapy. Advanced disease often follows a rapidly progressing course, and metastases to bone and lungs are common complications (1). To date, no clinical trial has been effective for treatment in the advanced or metastatic setting (9-11). Five-year overall survival (OS) in patients with NF1-MPNST is dismal at 20-50% (12). Numerous *in vivo* tumor models have been developed for preclinical studies, but there is limited genetic diversity modeled in these systems, which do not capture the full spectrum of genetic heterogeneity found in human NF1-MPNST. *TP53* loss, for example, is a common feature of the currently-used preclinical models, but is observed only in a minority of human MPNST(13, 14).

To address this critical problem, we have successfully generated a series of NF1-MPNST patient derived xenografts (PDX) that more accurately reflect the genetic heterogeneity seen in the human condition. Patient tumor tissue is implanted directly into immune-deficient mice; “*in vivo*” tumors are grown and serially passaged (15). These models better reflect the genomic and phenotypic heterogeneity of cancer (14). Here we present the development and comprehensive genomic characterization of eight NF1-MPNST PDX suitable as preclinical tools, to further study MPNST pathogenesis by identifying other drivers of transformation. In addition, we present the first use of single-cell RNA sequencing of NF1-MPNST (15, 16). Through characterization of these models, we identified a gain in the long arm of chromosome 8 (Chr8q) as a nearly universal event in MPNST. Further, Chr8q gain is associated with inferior outcomes in soft tissue sarcomas, which may contribute to the dismal overall survival of MPNST.

## Results and discussion

### Global characterization of PDX lines shows significant heterogeneity

Recently, PDX models have gained traction as a preclinical platform and a tool to study tumor heterogeneity. These humanized models represent a unique opportunity to study the characteristics of a large variety of tumor types, as early passage PDX lines retain a high degree of similarity to the parental tumor and allow study of tumor cell origin, biomarker identification, and treatment strategies (17-19). We generated and characterized eight NF1-MPNST PDX from eight corresponding biopsy-proven human NF1-MPNST between 2014 and 2019 at two institutions, Washington University and John Hopkins University (20). These lines represent the largest characterized set of NF1-MPNST PDX reported to date. The engraftment success rate was 50%. Whole exome and bulk RNA sequencing were performed at the McDonnell Genome Institute at Washington University. Clinical parameters are summarized in **Supplemental Table 1**. All tumors were high grade and greater than five centimeters. Seven patients were male, with a median age of onset of 34 years (range: 9-52 years). All PDX were histologically and morphologically comparable to the parental tumors, using hematoxylin and eosin (H&E), S100, Col4a, and Ki-67 staining **(Supplemental Fig. 1)**. Comprehensive analysis led to several important findings. First, we observed massive intertumoral heterogeneity within the cohort. Our somatic variant analysis identified an average of 80 high confidence single-nucleotide variants (SNV) and short indels per sample (range 13-99). Germline and common variants based on the 1000 genomes project (MAF > 0.05) were filtered out. *TP53* mutations occurred in 12.5% (1/8 pairs), *SUZ12* mutations in 62.5% (5/8 pairs), and the other 25% (2/8 pairs) had mutations in other cancer related pathways, such as DNA repair genes. The average mutation burden is 0.08 (range: 0.045-0.16) mutations/Mb (**Fig 1A)**.

**Fig. 1:**
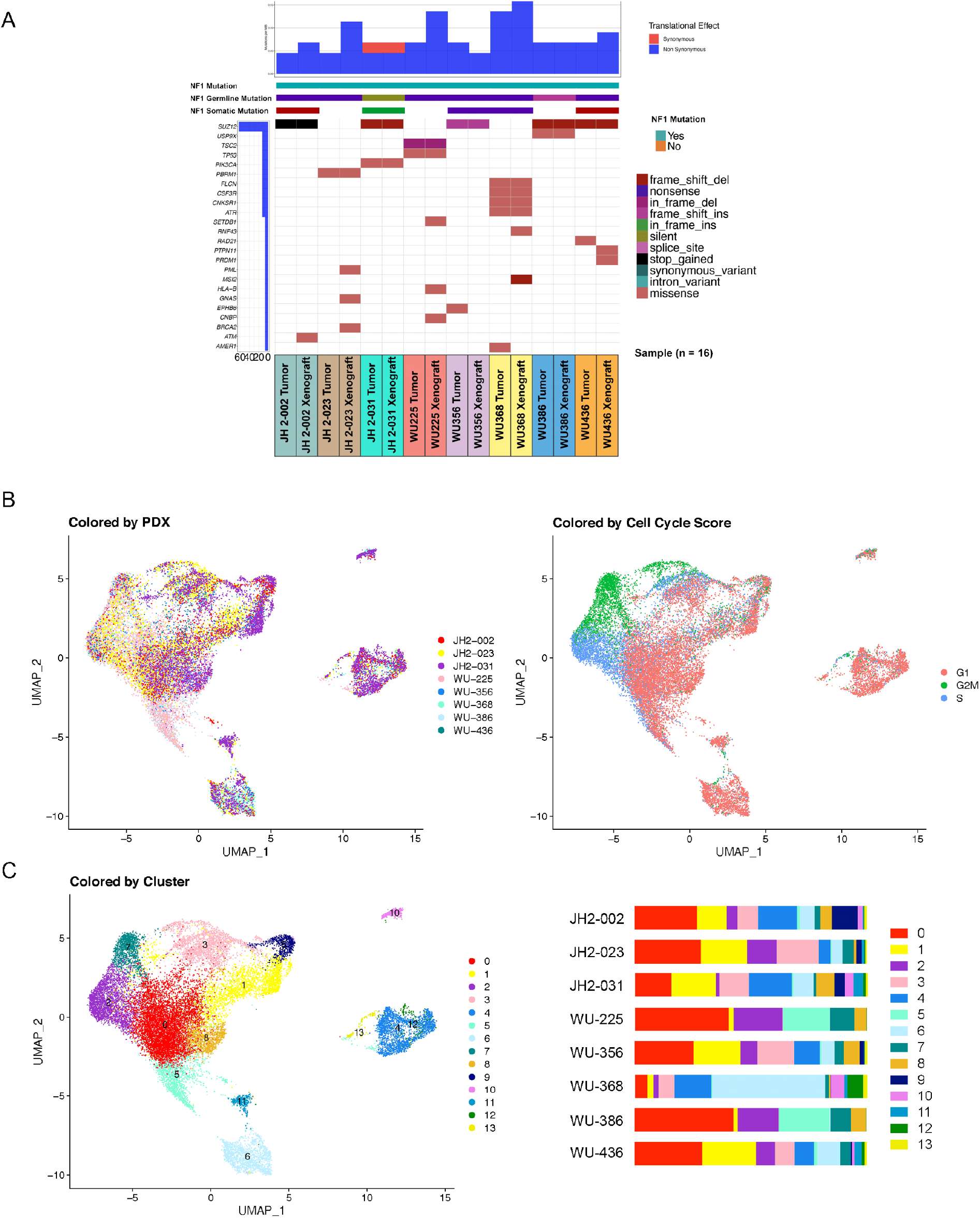
Whole exome sequencing and single cell composition of eight tumor-PDX pairs highlights inter-tumor heterogeneity. **A)** Somatic variants across samples. Distinct somatic variant signatures are appreciated in each tumor-PDX pair. **B)** UMAP of the eight MPNST PDX scRNAseq result shown as grouped by each PDX (left panel) or by cell cycle score (right panel). **C)** UMAP of all PDX shows each cluster and the bar plot demonstrates the inter-tumor heterogeneity across PDX as each line is composed of varying percentages of the different clusters.

To characterize the intra-tumoral heterogeneity at single-cell resolution, we performed droplet-based single cell RNA sequencing (scRNAseq) on 24055 cells from the eight MPNST PDX (**Supplemental Table 2**). Focusing on the expansion of human MPNST in our mouse model, downstream analysis used only sequencing reads that uniquely aligned to the human genome (GRCh38). Clustering of the scRNA-seq data revealed that cells from different PDX shared transcriptional similarities regardless of their cell cycle stages (**Fig. 1B**), separating the cells into 14 distinct clusters with each PDX contributing a unique distribution of cells to each cluster (**Fig. 1C; Supplemental Table 3**). Correlation of the VAF in the parental tumor versus the VAF in each PDX demonstrated the overall similarity of the PDX to its parental tumor. Interestingly, some variants showed a VAF difference, likely representing variants found predominantly in either the parental tumor or the PDX, suggestive that some clonal selection occurs upon engraftment (**Supplemental Fig. 2 left panel**). Scatter plots comparing the gene expression levels in the parental tumor versus the PDX showed an overall similarity, with a median correlation co-efficient of 0.79 (range: 0.66-0.92) (**Supplemental Fig. 2 right panel**). Hierarchical clustering by Euclidean distance and Z-scores were performed on all expressed genes in grouped parental tumor and PDX samples, further highlighting the intertumoral heterogeneity (**Supplemental Fig. 3**).

### CNV analysis by whole genome sequencing and single cell RNAseq demonstrated Chr8q gain in MPNST

In addition to coding sequence mutations and transcriptional expression, copy number variation (CNV) is another important source of genomic heterogeneity. We therefore analyzed the CNV profiles of the PDX, shown in **Fig. 2A**, and found remarkable similarities with the corresponding patient tumors. Notably, 80% of the cases showed a Chr8q gain. To determine if Chr8q gain could occur early in tumor development, we performed CNV analysis on seven PN, a precursor lesion of MPNST. Chr8q gain was not observed in any of the PN examined (**Fig. 2B**). We also found a high degree of aneuploidy in NF1-MPNST, whereas the PN samples harbored diploid genomes. DNA aneuploidy is known to be an independent risk factor, in addition to histologic grade and lymphovascular invasion, for decreased metastasis-free survival in sarcomas (21). Our data illustrate the difference in CNV between benign precursor lesions and the high-grade sarcoma after malignant transformation.

**Fig. 2:**
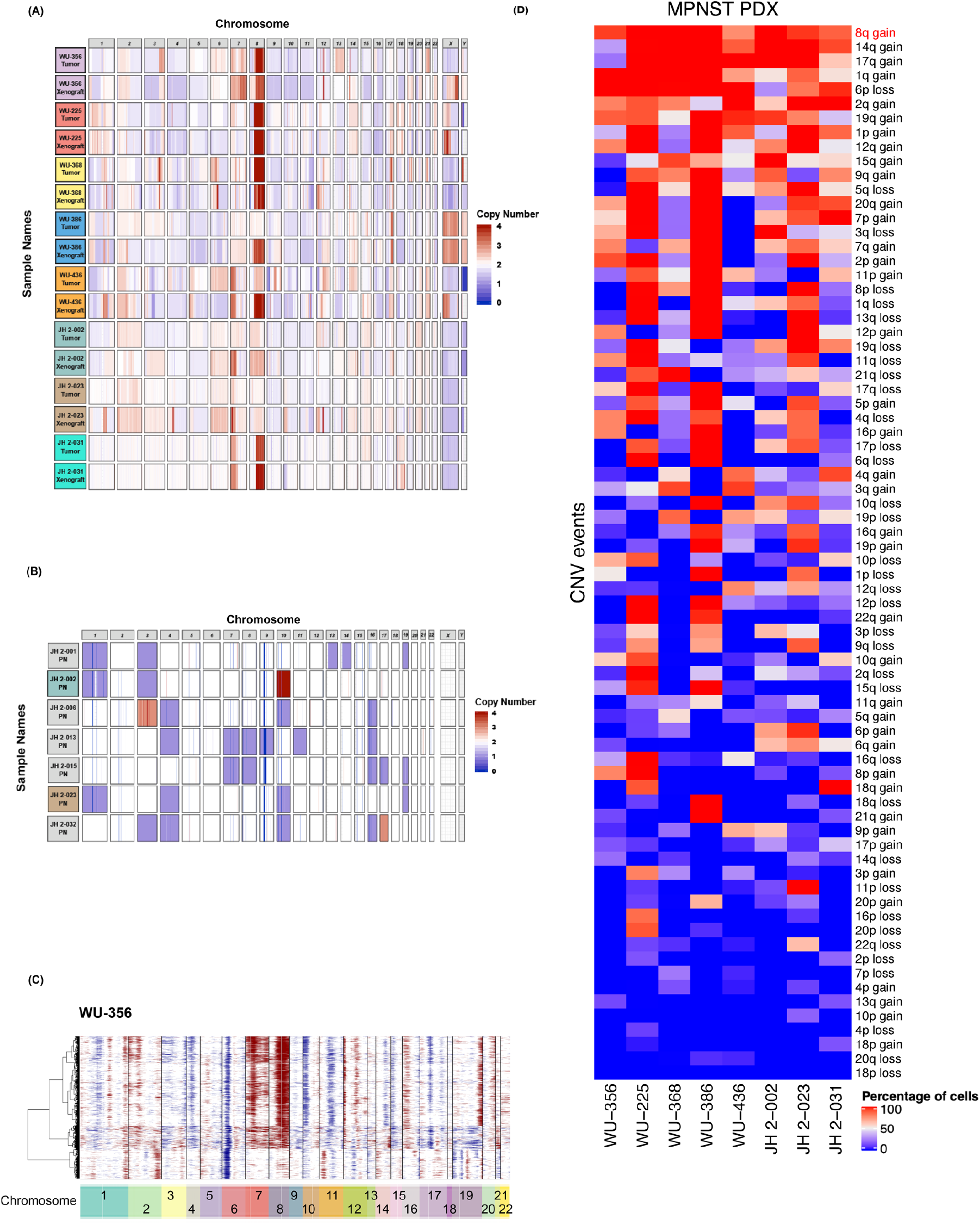
Chr8q gain is the most prevalent copy number variation in MPNST PDX. **A)** Copy number variation (CNV) plot of all eight MPNST tumor-PDX pairs. **B)** CNV plot of all seven PN samples. **C)** Representative CNV heatmap with hierarchical clustering of results from inferCNV analysis of scRNAseq result of WU-356. **D)** Summary heatmap of all large scale CNV events detected by inferCNV analysis of scRNAseq data of all eight MPNST PDX. Chr8q gain is the most prevalent CNV event among the 74 detected events.

The presence of CNV allowed us an opportunity to examine the clonal structure of each PDX generated with single cell data using the inferCNV method (22, 23). As expected, all PDX exhibited Chr8q gain in all or a portion of the tumor cells (**Fig. 2C, Supplemental Fig. 4**). A summary of each possible CNV event and the percentage of cells exhibiting these CNV events confirmed that Chr8q gain was the most prevalent and dominant CNV, present in >90% of cells (**Fig. 2D, Supplemental Table 4**). These data illustrate the first use of single cell sequencing in MPNST and demonstrate that Chr8q gain is an early, and highly recurrent CNV event among the 74 events detected in all eight NF1-MPNST.

Our analysis also revealed an under-appreciated complexity of clonal evolution of MPNST PDX in the mouse model. For instance, in WU-356, while Chr8q gain was a prominent event, along with 1q gain and 6p loss, we also appreciated the branching and evolution of subclone CNV (e.g., 11p gain, 12q loss, 19p loss) in 21% of all cells (**Fig. 2C and Fig. 3A**). Distinct patterns of genomic evolution shown by subclonal CNV events in each PDX indicate their evolutionary complexity in our model (**Fig. 3**) as well as in the primary tumors. While previous studies showed every chromosome can be involved in numerical and/or structural aberrations in MPNST (24), one study reported a Chr8 gain accompanied by alterations in several other chromosomes by array CGH (25). Our results specifically support Chr8q gain and suggest that Chr8q gain is important for high-grade transformation into MPNST.

**Fig. 3:**
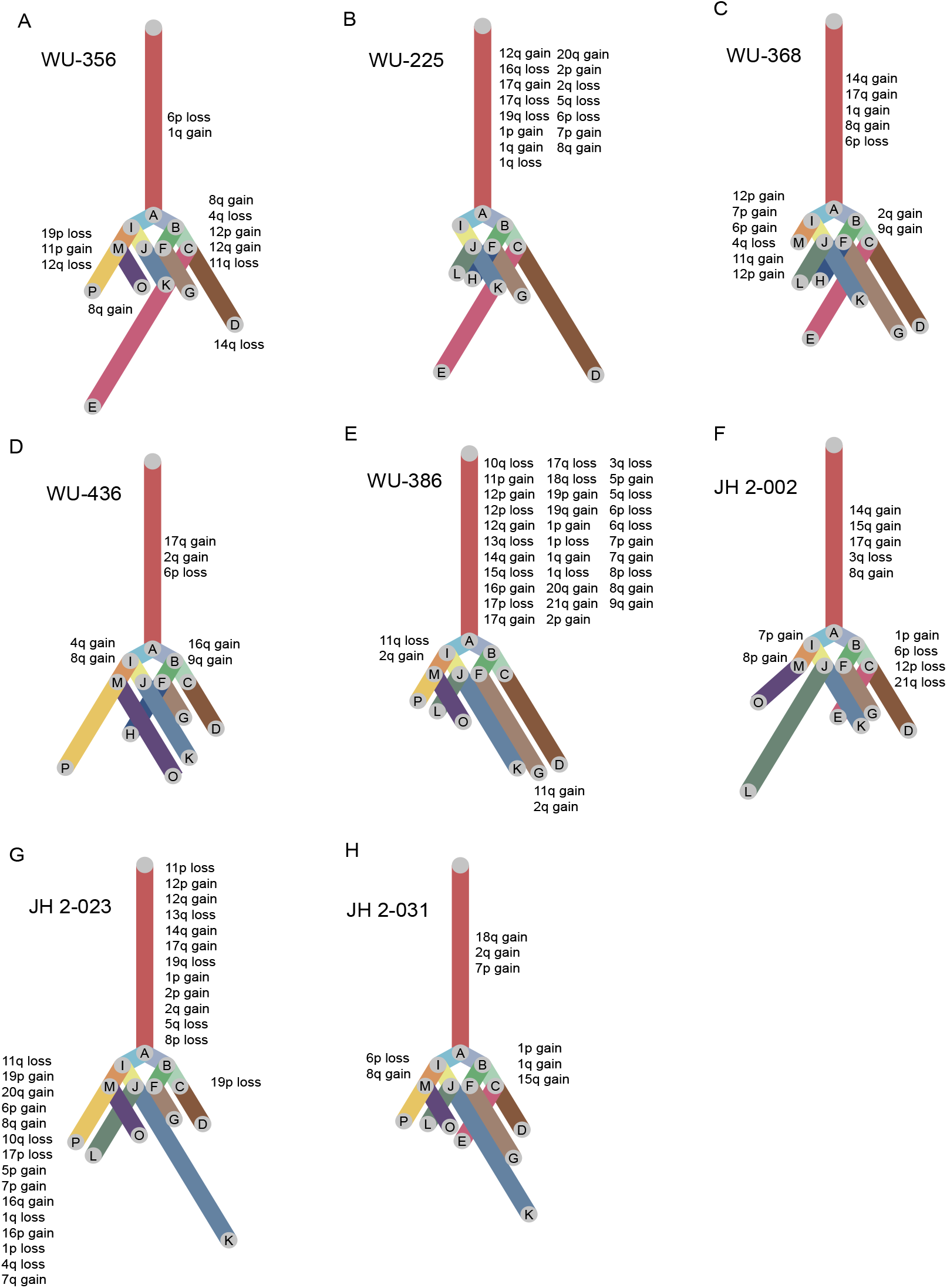
Clonal analysis of inferred CNV from scRNAseq highlight complexity in clonal evolution and depict Chr8q gain as an early CNV in MPNST-PDX. **A-H)** Clonality tree of each of the eight MPNST PDX. Lengths of the branches are scaled to the percentage of cells present in subclone of the corresponding CNV event.

### Chr8q gain is associated with MPNST transformation and poor overall survival in soft tissue sarcomas

To further validate the finding of Chr8q gain, we utilized fluorescence *in situ* hybridization (26) which permits the highly sensitive investigation of numerical chromosomal anomalies and genetic alterations on an individual cell basis within a region of interest (ROI) (**Fig. 4A**) (27). Chr8 spans about 145 million base pairs and 16% of its genes are implicated in cancer (28). **Fig 4B** depicts cancer-related genes in the area of Chr8q gain. Chr8 alterations are implicated in multiple malignancies, including translocations involving *c-myc* in Burkitt lymphoma (29). Additionally, certain solid tumors show extra copies of chr8, such as desmoid fibromatosis (30) and lipoblastoma (31, 32). Centromeric probe Chr8 FISH was performed on the eight PDX models, eight corresponding patient tumors, as well as ten other NF1-MPNST cases, with 200 cells counted per case. All but one PDX showed Chr8 gain, defined as three or more copies, in >50% of cells (7/8). Of the corresponding parental tumors, Chr8 gain was observed in five cases. However, all of the other cases (both PDX and parental tumors) demonstrated Chr8 gain in at least 10% of cells (**Fig 4A**). These data suggest a selective advantage for cells with Chr8 gain during the engraftment process. We also performed FISH on ten additional NF1-MPNST cases as a validation cohort. Similarly, in this cohort, eight out of ten NF1-MPNST had Chr8 gains in >50% of cells, and in the remaining two, Chr8 gains were present in at least 10% of cells. These findings strongly support the notion that Chr8 gain is a nearly universal event in MPNST.

**Fig. 4:**
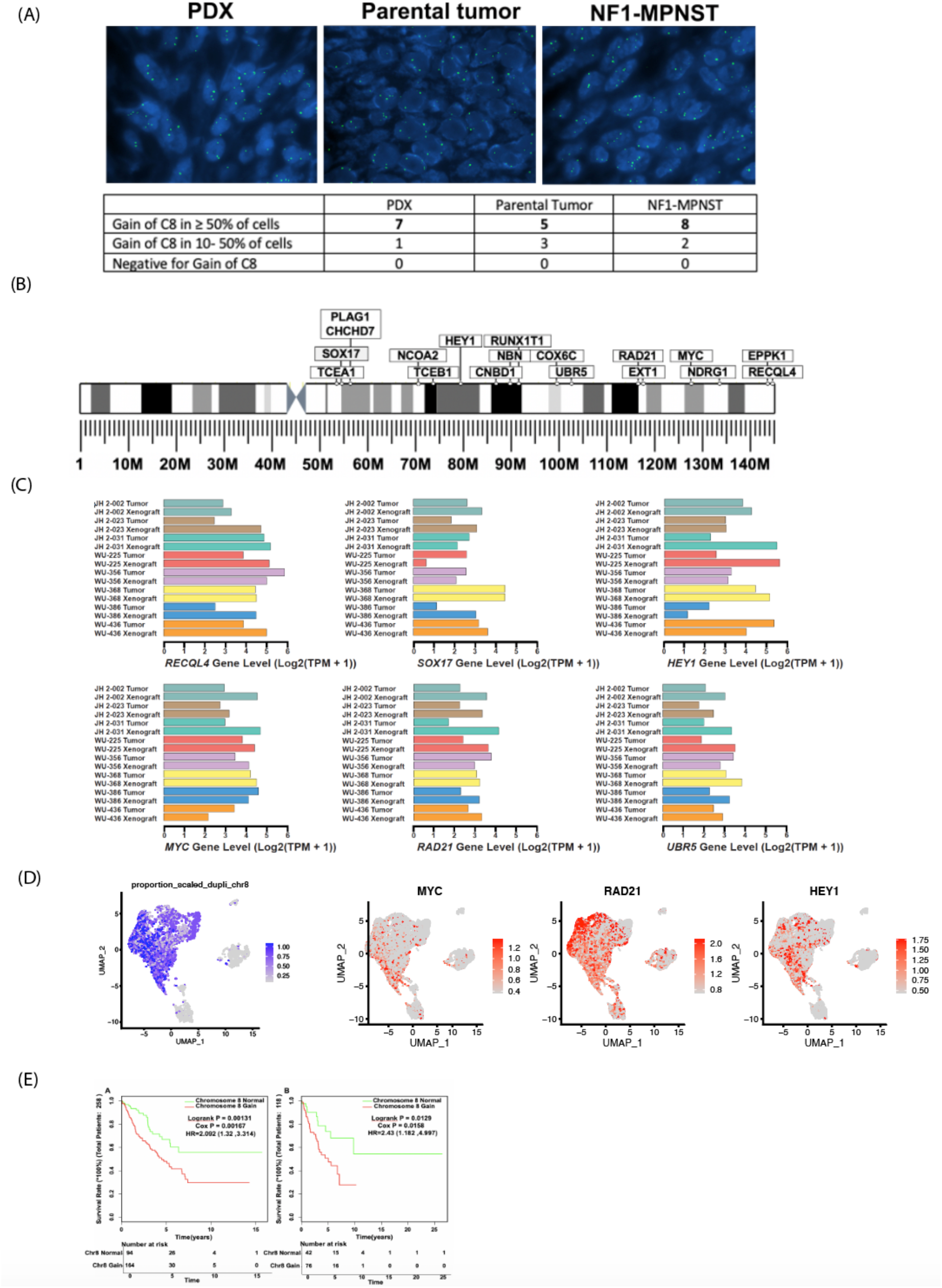
Chr8 gain is associated with MPNST transformation and inferior survival. **A)** Representative photographs of fluorescence *in situ* hybridization (26) signals per nucleus of PDX, parental tumor and control NF1-MPNST. Table below summarizes the results of the analysis. **B)** Important cancer-related genes on Chr8q. **C)** Expression levels of the top 6 genes in each PDX-tumor pair. **D)** UMAP of PDXs highlighting the existence of Chr8q gain, expression of *HEY1, MYC* and *RAD21*. **E)** Kaplan-Meir curves showing the association of Chr8q gain with inferior overall survival in patients with soft tissue sarcomas.

We next investigated the specific cancer-related genes located on Chr8q (**Fig. 4B**). Among these, *c-myc* (8q24) has been studied in a variety of cancers and known to be a cell cycle regulator, important for neoplastic transformation (33). Reuss and colleagues (34) showed that MPNST cell lines with complete *NF1* deficiency were sensitive to TRAIL-induced cell death that was critically dependent on *c-myc*. Thus, targeting dysregulated *c-myc* expression using BRD4 inhibitors may represent a therapeutic opportunity in NF1-associated MPNST (35). There are several other cancer-related genes on Chr8q including *UBR5* and *RAD21*, and we continue to investigate their potential role in MPNST tumorigenesis. Additionally, Chr8 gains have been described in Ewing sarcoma and several pediatric soft tissue sarcomas (36-38), and therefore may be a common founder event in sarcoma malignant transformation. We interrogated the expression levels of genes in this area of Chr8q using our bulk RNAseq data. The six genes with the highest expression are shown in **Fig. 4C**. The expression levels of the other genes were more variable and at much lower levels making them less likely candidates to play a role in progression of MPNST (**Supplemental Fig 5**). Next, comparing the cells with Chr8q gain to cells without Chr8q gain using the results of inferCNV, we obtained a list of significantly highly expressed genes, including *HEY1, MYC and RAD21* that were also revealed by bulk RNAseq (**Fig. 4, Supplemental Table 5**). Finally, using the cancer genome atlas (39) database, we analyzed the correlation between Chr8q gain and overall survival, and found that Chr8q gain was associated with inferior overall survival in two soft tissue sarcomas datasets (39) (**Fig. 4D** p=0.0013, p=0.0129). As every MPNST examined in our analysis demonstrated Chr8q gain, this association may be, at least in part, accountable for the poor survival seen in MPNST patients.

Through comprehensive characterization of a set of eight NF1-MPNST primary tumors and corresponding PDX models, and the first single-cell characterization of MPNST, our work highlights the complex inter- and intra-tumor heterogeneity found in MPNST. Our findings demonstrate that Chr8q gain is a common event in MPNST pathogenesis and a putative driver of malignant transformation. Future studies, conducted through multi-institutional collaboration, will be aimed at determining the strength of the correlation between CNV and clinical outcomes in a large subset of MPNST, as well as determination of the specific genes at this locus that are critical for MPNST progression.

## Data Availability

SYNAPSE repository: doi:10.7303/syn11638893

https://www.synapse.org/#!Synapse:syn11638893

## Author contributions

ACH obtained funding, conceptualized the research, designed and analyzed experiments, and edited the manuscript. JS, CAP, and SL aided in research conceptualization and experimental design and edited the manuscript. CD, CIM, ZZ, and Xiy Z designed experiments, analyzed data, and wrote the manuscript. Xia Z, KP, and TP generated PDX and edited the manuscript. CM, HX, XW, KY, JM, SG, YW, AG, PJ, SD, and HB performed experiments, analyzed data, and edited the manuscript.

## Acknowledgments

This work was funded by grants from the Neurofibromatosis Translational Acceleration Program (NTAP, to A.C.H. and C.A.P.). The NF Research Initiative (to A.C.H.), and the St. Louis Men’s Group Against Cancer (to A.C.H.)

