## Supplemental data and methods for "Chromosome 8 gain is associated with high-grade transformation in MPNST"

**Supplementary data**

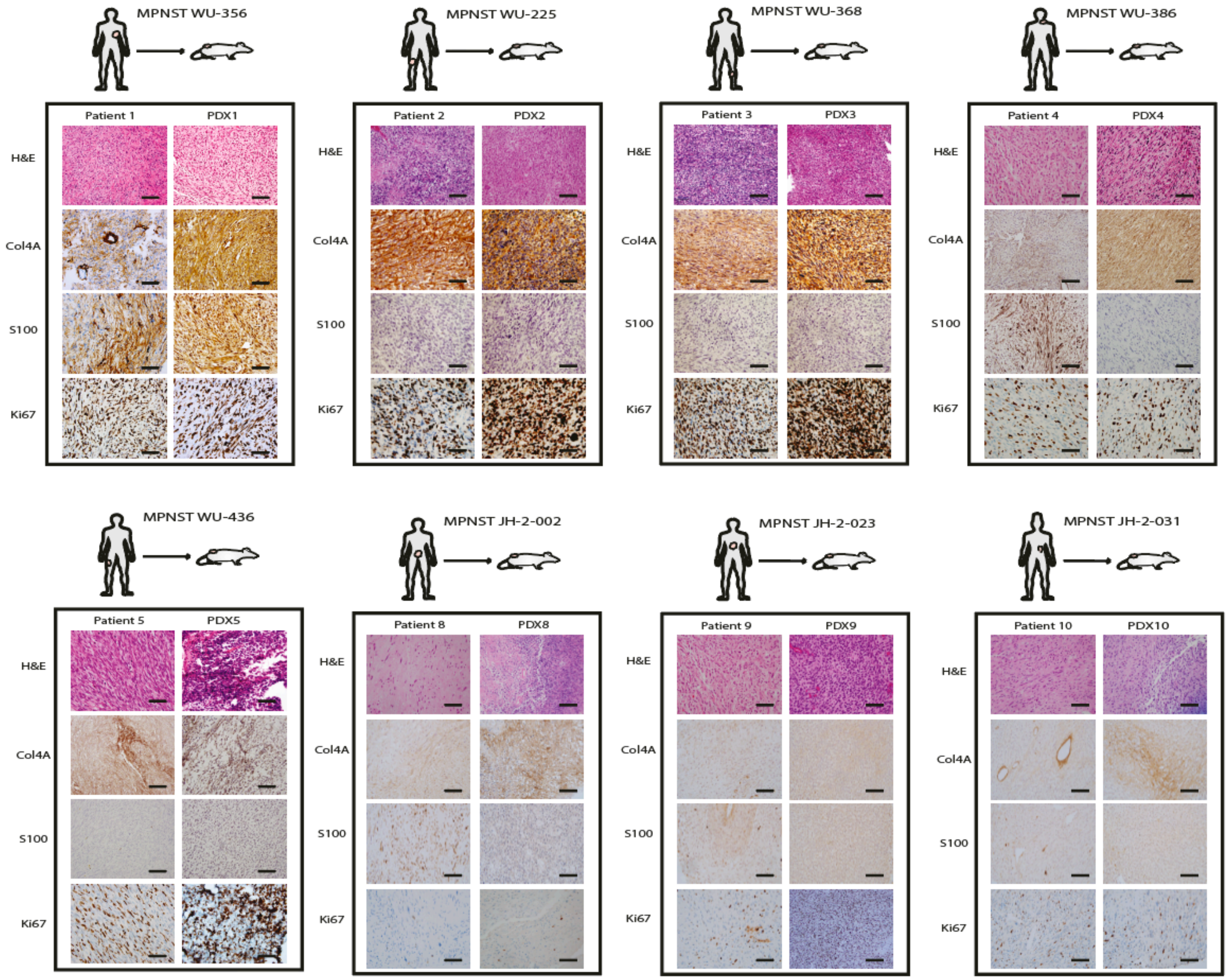


**Supplemental Fig. 1**: **Histological Comparison of tumor-of-origin to patient-derived-xenograft (PDX).** immunoreactivity for H&E, Col4A, S100, and Ki67 in patient tumors of origin and corresponding PDX tumors with schematic depicting the site of the original tumor in the patient. Scale bar = 20um.

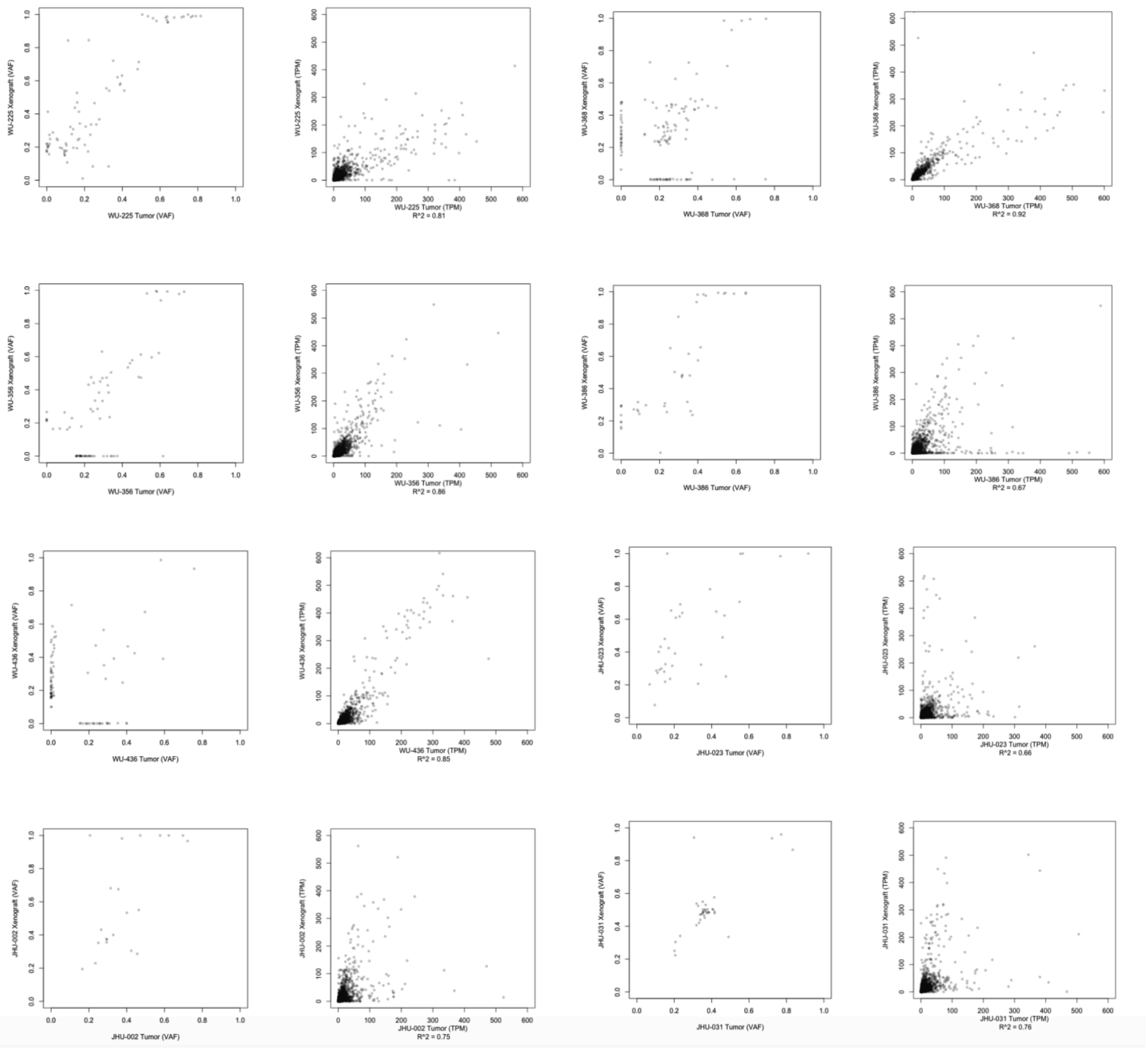


**Supplemental Fig. 2: Scatter plots displaying the correlation between tumor of origin and PDX DNA.**
Left: Tumor of origin v. patient derived xenograft (PDX) variant allele frequency (VAF) plots. Variance is in relation to the identity line, therefore, points along axis reflect evidence of heterogeneity. Right: Simple X/Y scatterplots showing raw counts gene expression levels of each tumor compared to its respective xenograft. The correlation coefficients (on a scale of 0 to 1) are included underneath.

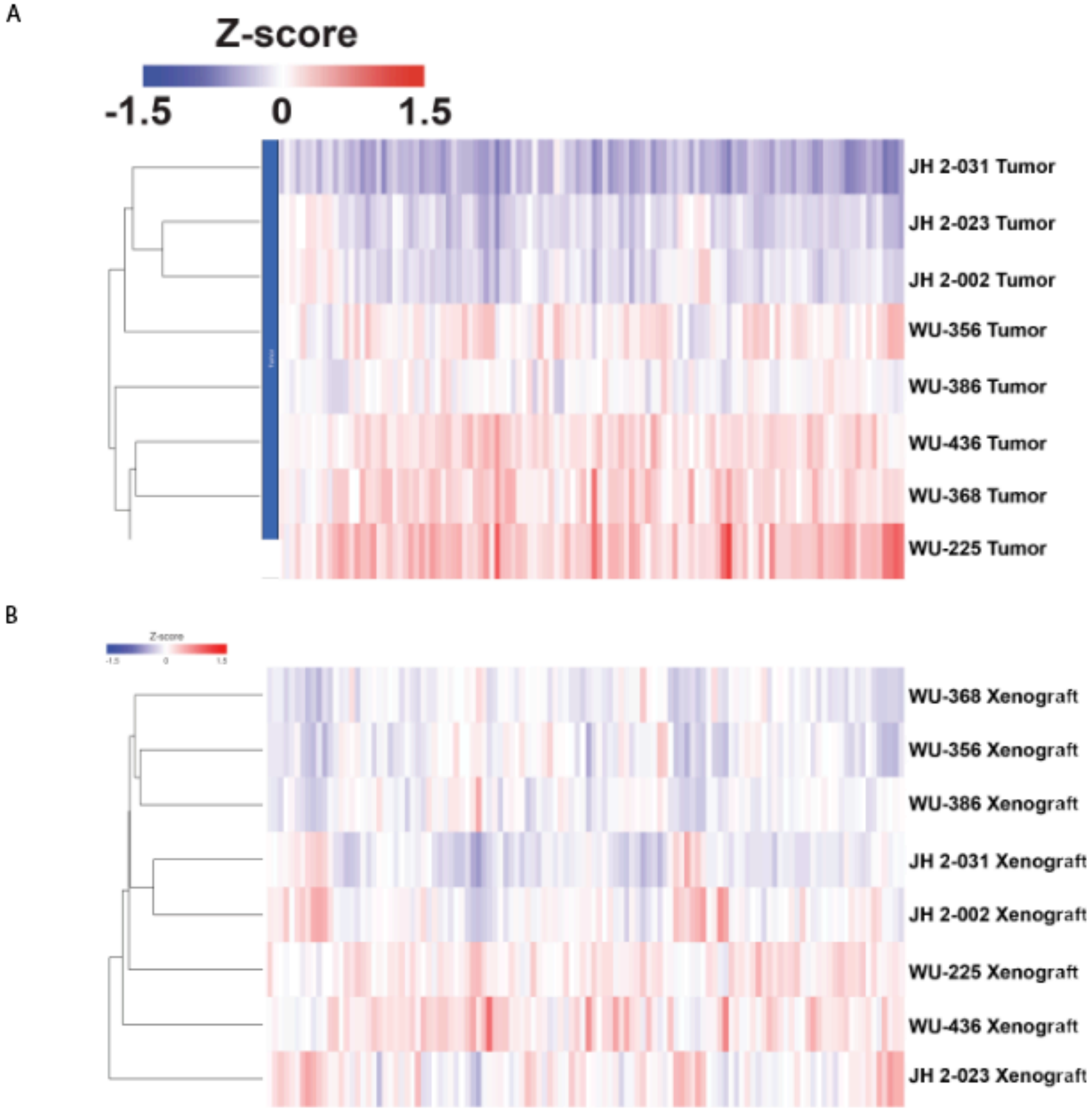


**Supplemental Fig. 3:**
A) Heatmap of eight parental tumor samples with list of all genes. B) Heatmap of eight PDX samples with list of all genes. Both rows and columns of heatmaps are clustered using Euclidean distance and average linkage

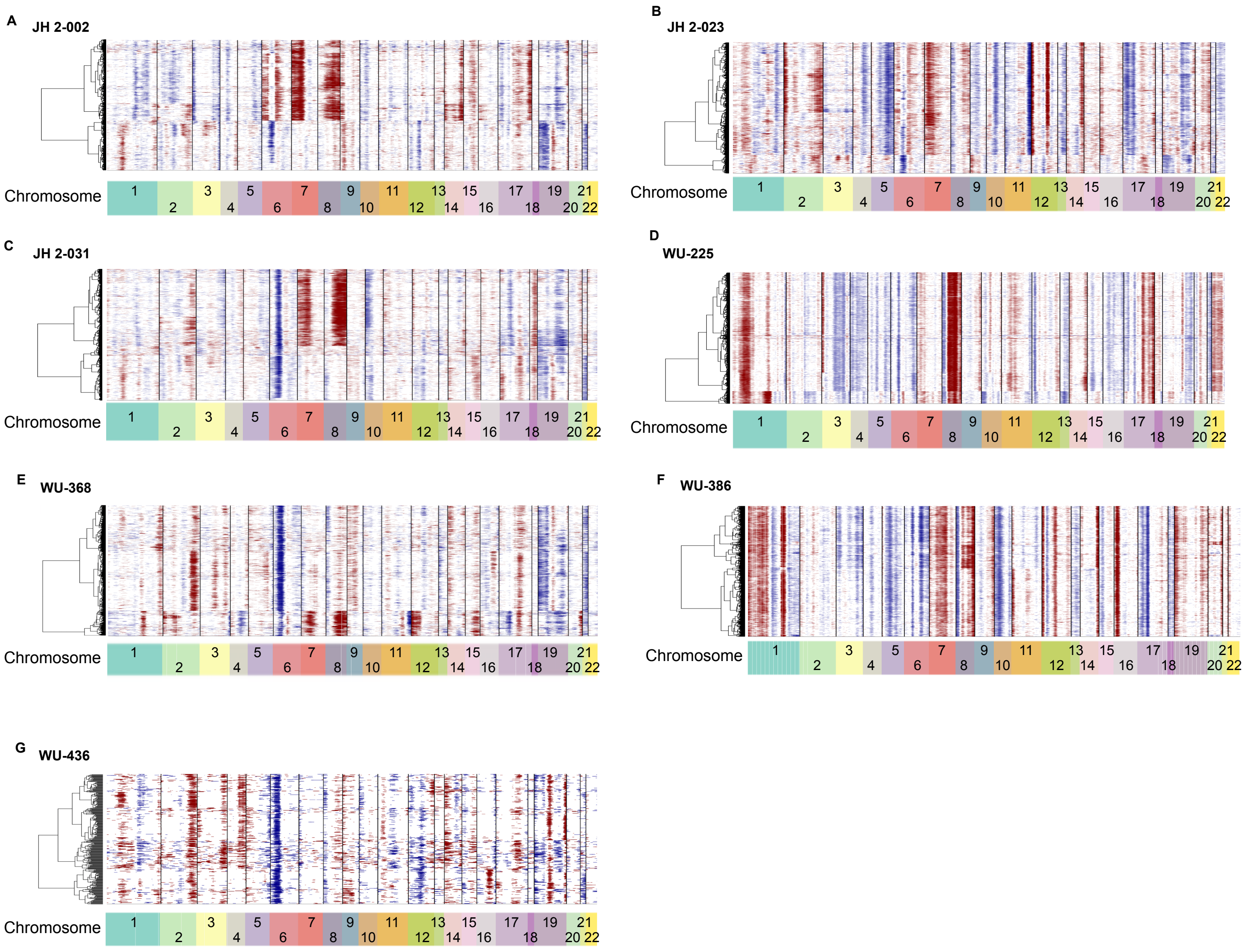


**Supplemental Figure 4: Gain of chromosome 8 is common in MPNST PDX.**
**A-G)** copy number variation (CNV) heatmap with hierarchical clustering of each MPNST PDX showing results of the inferCNV analysis.

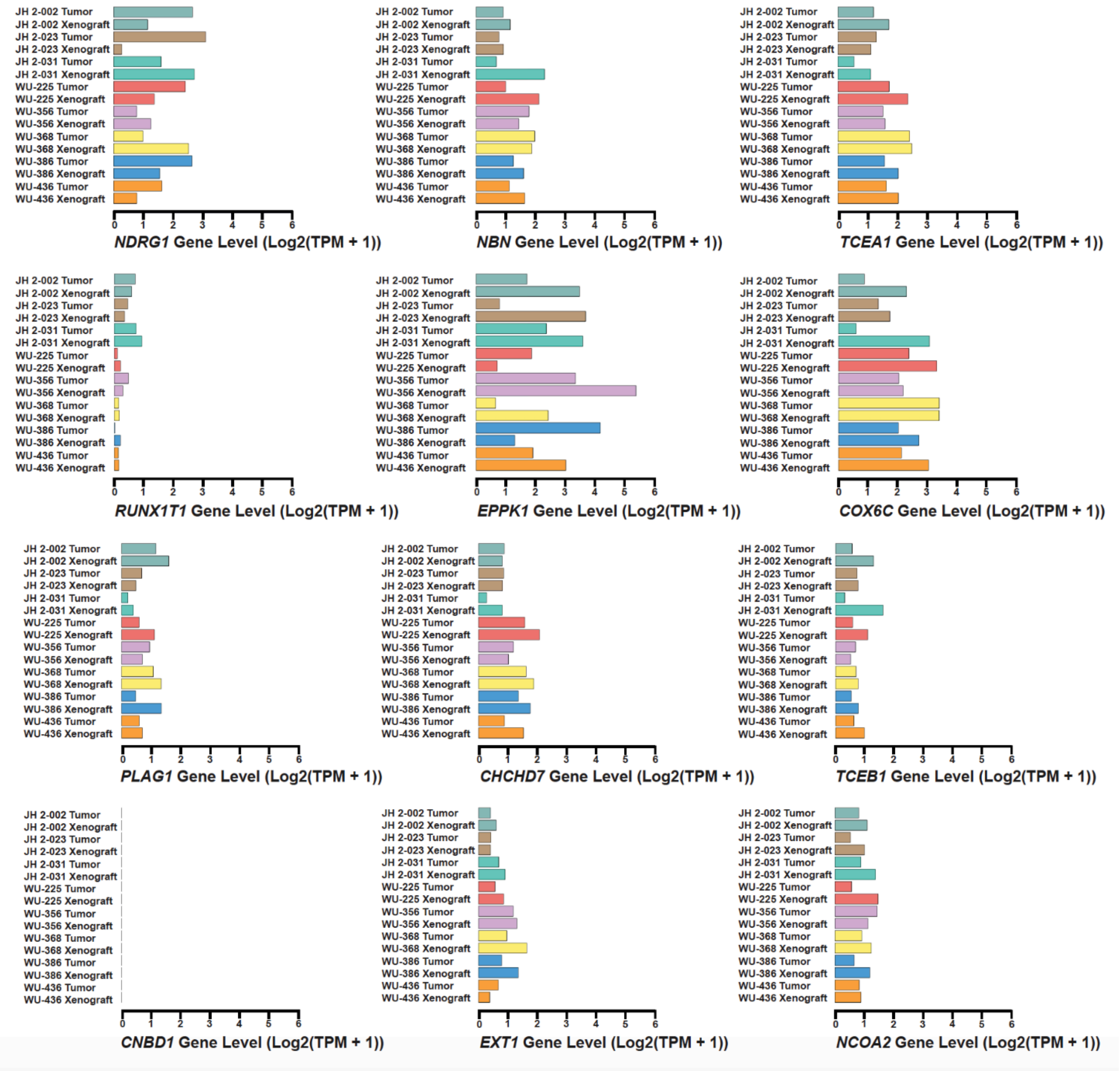


**Supplemental Fig. 5: Expression of additional cancer-related genes on Chromosome 8**
The barplots represent the RNA expression of additional cancer-related genes on chromosome 8 in our cohort.

Tables

| Tumor ID | Age range ~5-10 years | Sex | NF1 status | MPNST | Grade | Location | Size | Clinical Status |
|---|---|---|---|---|---|---|---|---|
| **WU-356** | 30ies | M | NF1 | Primary | High | Mediastinum | 12.6 | Deceased |
| **WU-225** | 40ies | M | NF1 | Primary | High | Thigh | 22 | Deceased |
| **WU-368** | 50ies | F | NF1 | Primary | High | Calf | 13.5 | NED |
| **WU-386** | 30ies | M | NF1 | Primary | High | Neck | 7 | NED |
| **WU-436** | 40ies | M | NF1 | Primary | High | Thigh | 15.8 | NED |
| **JH 2-002** | Teens | M | NF1 | Primary | High | Pelvis | 5.8 | NED |
| **JH 2-023** | 20ies | M | NF1 | Primary | High | Paraspinal | 6 | NED |
| **JH 2-031** | Teens | M | NF1 | Primary | High | Retroperitoneal | 10 | Deceased |

**Supplemental Table 1: Patient characteristics**
Overall, eight PDX were generated from eight NF1-patients with high-grade MPNST. The table displays the patient clinical characteristics used for the PDX models. NED = No evidence of disease

| Supplemental Table 2: Single-cell RNA sequencing QC | | | | |
|---|---|---|---|---|
| PDX ID | Total Cells Captured | Mean reads per cell | Median genes per cell | Cell count-GRCh38 |
| WU-386 | 3192 | 93371 | 4061 | 3106 |
| WU-225 | 4976 | 99624 | 5451 | 4948 |
| WU-368 | 5268 | 125664 | 976 | 1558 |
| WU-356 | 1968 | 114580 | 784 | 1644 |
| WU-436 | 2249 | 1265194 | 670 | 316 |
| JH2-023 | 4026 | 32401 | 2134 | 4590 |
| JH2-002 | 2390 | 75224 | 1812 | 2279 |
| JH2-031 | 6582 | 89513 | 1255 | 5614 |

**Supplemental Table 2: Single-cell RNA sequencing QC**

| | JH2-002 | JH2-023 | JH2-031 | WU-225 | WU-356 | WU-368 | WU-386 | WU-436 |
|---|---|---|---|---|---|---|---|---|
| 0 | 0.26853883 | 0.28540305 | 0.15817599 | 0.40602264 | 0.25364964 | 0.05455712 | 0.42691565 | 0.29113924 |
| 1 | 0.12856516 | 0.19978214 | 0.19059494 | 0.02081649 | 0.20194647 | 0.02759949 | 0.01835158 | 0.23101266 |
| 2 | 0.04651163 | 0.12636166 | 0.01531885 | 0.21059014 | 0.0729927 | 0.02053915 | 0.17611075 | 0.08227848 |
| 3 | 0.08951294 | 0.18126362 | 0.12771642 | 0.00181892 | 0.15632603 | 0.06931964 | 0.00128783 | 0.08544304 |
| 4 | 0.1667398 | 0.05141612 | 0.18453865 | 0 | 0.1107056 | 0.15789474 | 0 | 0.08227848 |
| 5 | 0.01140851 | 0.00457516 | 0.00552191 | 0.20230396 | 0.00790754 | 0.00449294 | 0.21796523 | 0.01265823 |
| 6 | 0.06537955 | 0.04662309 | 0.08656929 | 0.00080841 | 0.05778589 | 0.48395379 | 0.00450741 | 0.10126582 |
| 7 | 0.02281703 | 0.04901961 | 0.01086569 | 0.10448666 | 0.03832117 | 0.01797176 | 0.08757244 | 0.0443038 |
| 8 | 0.05221588 | 0.01176471 | 0.07908799 | 0.05234438 | 0.06812652 | 0.00577664 | 0.06535737 | 0 |
| 9 | 0.11057481 | 0.02287582 | 0.04453153 | 0 | 0.01946472 | 0.0012837 | 0 | 0.00632911 |
| 10 | 0.02062308 | 0 | 0.03758461 | 0 | 0 | 0.05840822 | 0 | 0.00949367 |
| 11 | 0.00482668 | 0.01132898 | 0.04061275 | 0.0004042 | 0.00182482 | 0.01412067 | 0.00032196 | 0.03164557 |
| 12 | 0.00263273 | 0.00326797 | 0.0122907 | 0.0004042 | 0.00121655 | 0.0661104 | 0.00160979 | 0.01265823 |
| 13 | 0.00965336 | 0.00631808 | 0.00659067 | 0 | 0.00973236 | 0.01797176 | 0 | 0.00949367 |

**Supplemental Table 3: Distribution of cell clusters among each PDX**

| | WU-356 | WU-225 | WU-368 | WU-386 | WU-436 | JH 2-002 | JH 2-023 | JH 2-031 |
|---|---|---|---|---|---|---|---|---|
| **8q gain** | 94.6570397 | 100 | 100 | 100 | 77.4193548 | 100 | 95.8634953 | 87.9599306 |
| **14q gain** | 30.4693141 | 99.9595469 | 100 | 100 | 89.0322581 | 100 | 100 | 93.0263918 |
| **17q gain** | 21.1552347 | 100 | 100 | 100 | 100 | 100 | 100 | 61.452514 |
| **1q gain** | 100 | 100 | 100 | 100 | 72.9032258 | 53.5601764 | 87.745605 | 54.4789058 |
| **6p loss** | 100 | 100 | 99.9354839 | 100 | 100 | 27.4102079 | 82.9886246 | 97.3030245 |
| **2q gain** | 80.433213 | 90.6148867 | 79.9354839 | 43.0560052 | 100 | 60.3024575 | 100 | 100 |
| **19q gain** | 88.8086643 | 90.5744337 | 51.483871 | 100 | 97.7419355 | 95.5891619 | 79.9120993 | 39.7418609 |
| **1p gain** | 36.534296 | 100 | 24.7741935 | 100 | 89.0322581 | 38.1222432 | 100 | 86.630707 |
| **12q gain** | 79.3501805 | 100 | 20 | 100 | 45.483871 | 79.7731569 | 100 | 48.2180697 |
| **15q gain** | 7.6534296 | 46.6019417 | 93.8064516 | 69.9579152 | 50.6451613 | 100 | 52.9472596 | 55.9815065 |
| **9q gain** | 0 | 90.5744337 | 80 | 100 | 27.4193548 | 89.0989288 | 13.081696 | 78.2893469 |
| **5q loss** | 1.37184116 | 100 | 55.2903226 | 100 | 53.8709677 | 73.7870195 | 100 | 53.8624542 |
| **20q gain** | 71.1913357 | 100 | 20 | 100 | 0 | 24.1965974 | 95.8634953 | 92.1017145 |
| **7p gain** | 56.7509025 | 100 | 20.0645161 | 100 | 0 | 64.4612476 | 91.0548087 | 100 |
| **3q loss** | 68.1588448 | 99.9595469 | 20 | 100 | 0 | 100 | 35.9358842 | 45.5210942 |
| **7q gain** | 79.3501805 | 9.42556634 | 69.1612903 | 100 | 0 | 64.4612476 | 86.1685626 | 58.8903872 |
| **2p gain** | 91.8411552 | 100 | 26.1290323 | 100 | 0 | 20.2268431 | 100 | 35.5422847 |
| **11p gain** | 19.2779783 | 90.6148867 | 49.0967742 | 100 | 65.1612903 | 22.9363579 | 0 | 64.7466769 |
| **8p loss** | 0 | 99.9595469 | 73.8064516 | 100 | 5.16129032 | 0 | 100 | 20.7281834 |
| **1q loss** | 0 | 100 | 0 | 100 | 45.483871 | 61.8777568 | 86.918304 | 13.369293 |
| **13q loss** | 5.34296029 | 99.9595469 | 35.4193548 | 100 | 2.25806452 | 0 | 100 | 6.97360817 |
| **12p gain** | 79.3501805 | 0.04045307 | 26.1935484 | 100 | 0 | 0 | 100 | 50.9921017 |
| **19q loss** | 17.9061372 | 100 | 48.4516129 | 0 | 21.6129032 | 73.7870195 | 100 | 79.2718166 |
| **11q loss** | 76.534296 | 99.9595469 | 20.0645161 | 43.0560052 | 23.5483871 | 24.7637051 | 94.131334 | 0 |
| **21q loss** | 5.34296029 | 90.5744337 | 99.9354839 | 0 | 27.4193548 | 35.5387524 | 60.8841779 | 32.7682527 |
| **17q loss** | 59.7833935 | 100 | 6.90322581 | 100 | 0 | 11.9092628 | 3.8262668 | 57.5611635 |
| **5p gain** | 13.9350181 | 90.5744337 | 31.6774194 | 100 | 46.4516129 | 0 | 92.78697 | 15.3149682 |
| **4q loss** | 79.3501805 | 99.9595469 | 20 | 91.9391389 | 0 | 60.0504096 | 86.1685626 | 0 |
| **16p gain** | 79.3501805 | 0.04045307 | 20 | 100 | 0 | 10.9010712 | 86.1685626 | 6.97360817 |
| **17p loss** | 0 | 90.6148867 | 13.1612903 | 100 | 0 | 61.8777568 | 89.9948294 | 13.369293 |
| **6q loss** | 0 | 99.9595469 | 0 | 100 | 0 | 0 | 0 | 20.7281834 |
| **4q gain** | 5.34296029 | 0.04045307 | 55.2903226 | 0 | 85.8064516 | 38.1222432 | 13.081696 | 93.0263918 |
| **3q gain** | 31.8411552 | 49.1100324 | 93.0967742 | 0 | 94.8387097 | 11.9092628 | 22.9317477 | 33.7507224 |
| **10q loss** | 0 | 19.7006473 | 0 | 100 | 0 | 77.189666 | 92.78697 | 20.7281834 |
| **19p loss** | 21.1552347 | 0 | 86.9032258 | 0 | 72.9032258 | 61.0586011 | 13.8314374 | 54.4789058 |
| **16q gain** | 5.34296029 | 0 | 22.3225806 | 97.6367757 | 27.4193548 | 9.3257719 | 89.2450879 | 8.34136005 |
| **19p gain** | 0 | 13.6731392 | 0 | 100 | 34.516129 | 0 | 96.9234747 | 6.97360817 |
| **10p loss** | 68.1588448 | 89.7249191 | 0 | 27.5493687 | 0 | 10.9010712 | 3.07652534 | 58.8903872 |

| | | | | | | | | |
|---|---|---|---|---|---|---|---|---|
| **1p loss** | 51.6245487 | 0 | 0 | 100 | 0 | 0 | 86.1685626 | 0 |
| **12q loss** | 6.71480144 | 7.36245955 | 0.06451613 | 0 | 78.3870968 | 38.1222432 | 70.7342296 | 28.2797149 |
| **12p loss** | 0 | 99.9595469 | 0 | 100 | 26.1290323 | 15.5009452 | 5.86866598 | 20.7281834 |
| **22q gain** | 0 | 99.9595469 | 0 | 97.6367757 | 0 | 0 | 0 | 6.97360817 |
| **3p loss** | 11.1913357 | 59.1423948 | 0.06451613 | 75.4613143 | 0 | 64.4612476 | 47.8024819 | 0 |
| **9q loss** | 8.15884477 | 76.0113269 | 0 | 71.090968 | 0 | 0 | 89.4002068 | 0 |
| **10q gain** | 59.7833935 | 92.6375405 | 0.06451613 | 0 | 4.19354839 | 22.9363579 | 0 | 58.8903872 |
| **2q loss** | 5.34296029 | 100 | 0 | 39.4626093 | 0 | 46.4398236 | 30.8428128 | 12.0400694 |
| **15q loss** | 30.7581227 | 97.9773463 | 0 | 100 | 5.16129032 | 0 | 0 | 0 |
| **11q gain** | 0 | 21.9660194 | 26.1290323 | 54.5807705 | 0 | 15.5009452 | 39.1158221 | 0 |
| **5q gain** | 4.40433213 | 9.38511327 | 55.2903226 | 0 | 13.2258065 | 11.9092628 | 3.07652534 | 25.4093624 |
| **6p gain** | 0 | 9.42556634 | 20.0645161 | 0 | 0 | 72.5897921 | 96.9234747 | 0 |
| **6q gain** | 11.1913357 | 0.04045307 | 0.06451613 | 0 | 0 | 64.4612476 | 76.9906929 | 51.916779 |
| **16q loss** | 31.8411552 | 100 | 6.19354839 | 2.36322434 | 51.9354839 | 2.58349086 | 7.21302999 | 0 |
| **8p gain** | 79.3501805 | 99.9595469 | 0.06451613 | 0 | 0 | 10.9010712 | 0 | 13.369293 |
| **18q gain** | 0 | 90.6148867 | 0.06451613 | 0 | 0 | 0 | 0 | 100 |
| **18q loss** | 0 | 9.38511327 | 0 | 100 | 0 | 0 | 17.0113754 | 0 |
| **21q gain** | 11.1913357 | 0.04045307 | 0 | 100 | 0 | 0 | 0 | 13.369293 |
| **9p gain** | 5.34296029 | 0 | 20.0645161 | 0 | 68.3870968 | 61.8777568 | 7.21302999 | 0 |
| **17p gain** | 13.9350181 | 7.36245955 | 0 | 0 | 5.16129032 | 28.7964713 | 5.86866598 | 39.7418609 |
| **14q loss** | 19.566787 | 0.04045307 | 6.90322581 | 0 | 0 | 0 | 22.1044467 | 6.97360817 |
| **3p gain** | 0 | 80.3398058 | 26.8387097 | 0 | 27.0967742 | 0 | 3.07652534 | 0 |
| **11p loss** | 0 | 7.40291262 | 0 | 0 | 2.25806452 | 11.9092628 | 100 | 0 |
| **20p gain** | 0 | 9.38511327 | 0.06451613 | 67.2709615 | 0 | 9.07372401 | 22.1044467 | 0 |
| **16p loss** | 0 | 85.3964401 | 0 | 0 | 0 | 0 | 7.21302999 | 0 |
| **20p loss** | 0 | 90.5744337 | 0 | 5.69763678 | 0 | 0 | 0 | 0 |
| **22q loss** | 0 | 10.315534 | 6.12903226 | 0 | 2.25806452 | 0 | 64.0641158 | 0 |
| **2p loss** | 0 | 13.7135922 | 0 | 0 | 0 | 0 | 0 | 18.0312079 |
| **7p loss** | 0 | 0 | 22.3225806 | 0 | 4.19354839 | 0 | 0 | 0 |
| **4p gain** | 0 | 0.04045307 | 13.1612903 | 0 | 2.25806452 | 0 | 9.85005171 | 0 |
| **13q gain** | 11.1913357 | 0.04045307 | 0 | 0 | 0 | 0 | 0 | 13.369293 |
| **10p gain** | 0 | 0 | 0 | 0 | 0 | 0 | 17.0113754 | 0 |
| **4p loss** | 0 | 9.38511327 | 0 | 0 | 0 | 0 | 0 | 0 |
| **18p gain** | 0 | 2.02265372 | 0 | 0 | 0 | 0 | 0 | 13.369293 |
| **20q loss** | 0 | 0 | 0.06451613 | 5.69763678 | 4.19354839 | 0 | 0 | 0 |
| **18p loss** | 0 | 0 | 0 | 0 | 0 | 0 | 0 | 0 |

**Supplemental Table 4: Detailed information of clonality calculation from inferCNV**

| | genes | p_val | avg_logFC | pct.1 | pct.2 | p_val_adj | chr |
|---|---|---|---|---|---|---|---|
| 137 | CD63 | 0 | 1.56849917 | 0.853 | 0.53 | 0 | chr12 |
| 180 | COL6A1 | 0 | 1.60851759 | 0.81 | 0.235 | 0 | chr21 |
| 206 | CTHRC1 | 0 | 1.78973995 | 0.837 | 0.313 | 0 | chr8 |
| 237 | DLK1 | 0 | 1.98257785 | 0.778 | 0.163 | 0 | chr14 |
| 251 | EEF1D | 0 | 0.98536258 | 0.918 | 0.641 | 0 | chr8 |
| 365 | IGF2 | 0 | 2.54555309 | 0.578 | 0.005 | 0 | chr11 |
| 503 | NEAT1 | 0 | 1.82845077 | 0.862 | 0.383 | 0 | chr11 |
| 523 | NUPR1 | 0 | 1.23828515 | 0.727 | 0.089 | 0 | chr16 |
| 71 | B2M | 5.65E-306 | 1.35108218 | 0.89 | 0.58 | 1.66E-301 | chr15 |
| 779 | TIMP1 | 4.27E-305 | 1.38515144 | 0.855 | 0.468 | 1.25E-300 | chrX |
| 772 | TAGLN2 | 4.39E-293 | 1.17018998 | 0.702 | 0.179 | 1.29E-288 | chr1 |
| 493 | NDUFA4 | 2.10E-290 | 0.99344943 | 0.867 | 0.505 | 6.15E-286 | chr7 |
| 323 | GNB2L1 | 4.74E-287 | 2.36400984 | 0.53 | 0.001 | 1.39E-282 | chr5 |
| 399 | LDHA | 6.80E-287 | 1.07809843 | 0.875 | 0.556 | 2.00E-282 | chr11 |
| 51 | ATP5E | 1.23E-286 | 1.74022421 | 0.53 | 0.001 | 3.59E-282 | chr20 |
| 60 | ATP5L | 2.21E-286 | 1.39552167 | 0.529 | 0.001 | 6.50E-282 | chr11 |
| 91 | C14orf2 | 2.51E-286 | 1.48228773 | 0.529 | 0.001 | 7.38E-282 | chr14 |
| 775 | TCEB2 | 3.28E-286 | 1.66695322 | 0.529 | 0.001 | 9.62E-282 | chr16 |
| 48 | ATP5B | 6.01E-286 | 1.36937571 | 0.528 | 0.001 | 1.76E-281 | chr12 |
| 66 | ATPIF1 | 6.55E-284 | 1.23247632 | 0.526 | 0.001 | 1.92E-279 | chr1 |
| 710 | SHFM1 | 1.74E-283 | 1.11380098 | 0.526 | 0.001 | 5.11E-279 | chr7 |
| 55 | ATP5G3 | 4.94E-283 | 1.23213217 | 0.524 | 0.001 | 1.45E-278 | chr2 |
| 58 | ATP5J | 5.22E-283 | 1.03121765 | 0.524 | 0.001 | 1.53E-278 | chr21 |
| 328 | GPX1 | 3.39E-282 | 1.19178617 | 0.524 | 0.001 | 9.94E-278 | chr3 |
| 61 | ATP5O | 4.76E-282 | 1.03948308 | 0.523 | 0.001 | 1.40E-277 | chr21 |
| 570 | PLD3 | 6.51E-281 | 1.08527458 | 0.767 | 0.299 | 1.91E-276 | chr19 |
| 200 | CRIP2 | 2.83E-280 | 0.72571445 | 0.607 | 0.056 | 8.32E-276 | chr14 |
| 774 | TCEB1 | 4.38E-280 | 1.10188505 | 0.521 | 0.001 | 1.29E-275 | chr8 |
| 576 | PLP2 | 7.84E-279 | 0.71195934 | 0.627 | 0.084 | 2.30E-274 | chrX |
| 47 | ATP5A1 | 1.25E-278 | 1.18747793 | 0.519 | 0.001 | 3.67E-274 | chr18 |
| 54 | ATP5G2 | 1.01E-276 | 0.98864548 | 0.516 | 0.001 | 2.96E-272 | chr12 |

| | | | | | | | |
|---|---|---|---|---|---|---|---|
| 50 | ATP5D | 1.07E-276 | 0.91859071 | 0.516 | 0.001 | 3.14E-272 | chr19 |
| 90 | C14orf166 | 3.06E-275 | 0.89135035 | 0.514 | 0.001 | 8.98E-271 | chr14 |
| 98 | C19orf43 | 1.59E-273 | 0.77616357 | 0.511 | 0.001 | 4.68E-269 | chr19 |
| 56 | ATP5H | 2.44E-272 | 0.85641502 | 0.51 | 0.001 | 7.17E-268 | chr17 |
| 588 | POSTN | 2.38E-271 | 1.38430018 | 0.7 | 0.14 | 6.97E-267 | chr13 |
| 132 | CD151 | 4.38E-269 | 0.73289773 | 0.697 | 0.154 | 1.29E-264 | chr11 |
| 856 | WBP5 | 5.35E-269 | 0.91818938 | 0.505 | 0.001 | 1.57E-264 | chrX |
| 505 | NGFRAP1 | 9.18E-269 | 0.90854731 | 0.505 | 0.001 | 2.69E-264 | chrX |
| 136 | CD59 | 1.20E-268 | 0.8403912 | 0.659 | 0.139 | 3.52E-264 | chr11 |
| 656 | RP11-14N7.2 | 2.65E-268 | 0.89827715 | 0.503 | 0 | 7.77E-264 | chr1 |
| 59 | ATP5J2 | 2.33E-267 | 0.83737858 | 0.503 | 0.001 | 6.83E-263 | chr7 |
| 349 | HN1 | 2.37E-267 | 1.27019371 | 0.503 | 0.001 | 6.95E-263 | chr17 |
| 510 | NPC2 | 1.67E-266 | 1.08450873 | 0.761 | 0.291 | 4.91E-262 | chr14 |
| 343 | HLA-B | 7.41E-266 | 1.27261371 | 0.846 | 0.431 | 2.17E-261 | chr6 |
| 53 | ATP5G1 | 1.97E-265 | 1.07347016 | 0.5 | 0.001 | 5.77E-261 | chr17 |
| 476 | MYEOV2 | 5.45E-265 | 0.70712602 | 0.499 | 0 | 1.60E-260 | chr2 |
| 582 | POLR2J3 | 8.57E-264 | 0.59523707 | 0.529 | 0.016 | 2.52E-259 | chr7 |
| 737 | SNHG7 | 2.59E-262 | 0.71114696 | 0.736 | 0.195 | 7.60E-258 | chr9 |
| 57 | ATP5I | 2.63E-262 | 0.79199311 | 0.496 | 0.001 | 7.70E-258 | chr4 |
| 361 | IER3 | 1.06E-261 | 0.92288238 | 0.636 | 0.095 | 3.12E-257 | chr6 |
| 622 | PTRF | 1.07E-261 | 0.81973884 | 0.494 | 0 | 3.15E-257 | chr17 |
| 638 | RBM3 | 2.17E-261 | 0.90801353 | 0.802 | 0.35 | 6.37E-257 | chrX |
| 321 | GLTSCR2 | 4.26E-261 | 0.7588206 | 0.494 | 0.001 | 1.25E-256 | chr19 |
| 843 | USMG5 | 3.31E-260 | 0.66542969 | 0.493 | 0.001 | 9.71E-256 | chr10 |
| 190 | COX6C | 1.01E-259 | 1.06899834 | 0.874 | 0.557 | 2.97E-255 | chr8 |
| 439 | MFAP4 | 4.39E-258 | 0.80728756 | 0.62 | 0.101 | 1.29E-253 | chr17 |
| 806 | TNFRSF12A | 4.45E-258 | 1.06988968 | 0.565 | 0.052 | 1.30E-253 | chr16 |
| 574 | PLOD1 | 8.35E-256 | 0.72667909 | 0.682 | 0.16 | 2.45E-251 | chr1 |
| 95 | C17orf89 | 3.65E-255 | 0.73925819 | 0.485 | 0 | 1.07E-250 | chr17 |
| 52 | ATP5F1 | 6.15E-255 | 0.82539951 | 0.485 | 0.001 | 1.81E-250 | chr1 |
| 222 | DCN | 2.21E-254 | 1.16206108 | 0.682 | 0.152 | 6.50E-250 | chr12 |
| 84 | C11orf31 | 1.26E-252 | 0.8289279 | 0.482 | 0.001 | 3.71E-248 | chr11 |
| 815 | TRAM1 | 2.52E-252 | 0.75098551 | 0.752 | 0.242 | 7.38E-248 | chr8 |
| 299 | FGFR1 | 6.06E-252 | 0.87494694 | 0.75 | 0.225 | 1.78E-247 | chr8 |
| 693 | SELM | 5.26E-251 | 0.68158388 | 0.479 | 0 | 1.54E-246 | chr22 |
| 78 | BNIP3 | 6.40E-251 | 0.98916595 | 0.738 | 0.201 | 1.88E-246 | chr10 |
| 836 | UFD1L | 3.88E-248 | 0.63434213 | 0.475 | 0.001 | 1.14E-243 | chr22 |
| 473 | MXRA8 | 8.35E-247 | 0.85653475 | 0.663 | 0.171 | 2.45E-242 | chr1 |
| 738 | SNHG8 | 8.49E-247 | 0.85542024 | 0.752 | 0.233 | 2.49E-242 | chr4 |
| 342 | HLA-A | 2.09E-246 | 1.11112142 | 0.834 | 0.398 | 6.13E-242 | chr6 |

| | | | | | | | |
|---|---|---|---|---|---|---|---|
| 699 | SEPW1 | 7.07E-245 | 0.60526995 | 0.471 | 0.001 | 2.07E-240 | chr19 |
| 751 | SPON2 | 1.44E-243 | 0.89543698 | 0.48 | 0.009 | 4.21E-239 | chr4 |
| 654 | RNH1 | 2.11E-243 | 0.62342584 | 0.694 | 0.173 | 6.19E-239 | chr11 |
| 49 | ATP5C1 | 1.49E-242 | 0.61402305 | 0.466 | 0 | 4.36E-238 | chr10 |
| 280 | FABP5 | 4.12E-242 | 1.47073091 | 0.663 | 0.178 | 1.21E-237 | chr8 |
| 284 | FAM195B | 7.67E-240 | 0.61166934 | 0.462 | 0 | 2.25E-235 | chr17 |
| 75 | BGN | 3.11E-239 | 0.84924806 | 0.677 | 0.137 | 9.11E-235 | chrX |
| 36 | APOA1BP | 1.72E-238 | 0.53299114 | 0.46 | 0 | 5.06E-234 | chr1 |
| 607 | PSAP | 4.71E-238 | 0.99836751 | 0.742 | 0.289 | 1.38E-233 | chr10 |
| 766 | STRA13 | 4.85E-238 | 0.6002639 | 0.46 | 0 | 1.42E-233 | chr17 |
| 356 | HTRA1 | 5.00E-237 | 0.85551869 | 0.735 | 0.216 | 1.47E-232 | chr10 |
| 403 | LHFP | 1.25E-236 | 0.59630002 | 0.459 | 0.001 | 3.66E-232 | chr13 |
| 553 | PGAM1 | 1.36E-236 | 0.77863388 | 0.792 | 0.34 | 3.99E-232 | chr10 |
| 695 | 15-Sep | 2.38E-235 | 0.50507908 | 0.456 | 0 | 6.98E-231 | chr1 |
| 113 | C7orf73 | 4.13E-235 | 0.47483222 | 0.456 | 0.001 | 1.21E-230 | chr7 |
| 787 | TMED9 | 6.58E-235 | 0.72820563 | 0.732 | 0.243 | 1.93E-230 | chr5 |
| 721 | SLC25A6 | 6.57E-234 | 0.88186497 | 0.848 | 0.44 | 1.93E-229 | chrX |
| 562 | PHLDA3 | 7.39E-234 | 0.56933144 | 0.554 | 0.064 | 2.17E-229 | chr1 |
| 615 | PSMG3 | 1.20E-233 | 0.54723477 | 0.645 | 0.137 | 3.51E-229 | chr7 |
| 662 | RPS17 | 2.53E-233 | 1.07552766 | 0.876 | 0.632 | 7.43E-229 | chr15 |
| 334 | GUK1 | 7.82E-232 | 0.79296907 | 0.816 | 0.364 | 2.30E-227 | chr1 |
| 181 | COL6A2 | 2.34E-230 | 1.52283938 | 0.77 | 0.31 | 6.87E-226 | chr21 |
| 840 | UQCRH | 3.18E-230 | 0.79526055 | 0.829 | 0.445 | 9.34E-226 | chr1 |
| 839 | UQCRFS1 | 5.13E-230 | 0.72769596 | 0.669 | 0.181 | 1.51E-225 | chr19 |
| 692 | SELK | 6.00E-228 | 0.46391356 | 0.446 | 0.001 | 1.76E-223 | chr3 |
| 105 | C20orf24 | 2.39E-227 | 0.48001645 | 0.445 | 0.001 | 7.02E-223 | chr20 |
| 713 | SHMT2 | 2.53E-227 | 0.52685666 | 0.625 | 0.118 | 7.43E-223 | chr12 |
| 853 | VIMP | 4.21E-227 | 0.50772878 | 0.445 | 0.001 | 1.24E-222 | chr15 |
| 858 | WBSCR22 | 7.01E-227 | 0.4288672 | 0.445 | 0.001 | 2.06E-222 | chr7 |
| 285 | FAM20C | 2.29E-226 | 0.49568964 | 0.541 | 0.057 | 6.72E-222 | chr7 |
| 845 | UTP11L | 2.94E-226 | 0.47049888 | 0.442 | 0 | 8.62E-222 | chr1 |
| 784 | TMED2 | 5.75E-226 | 0.66866357 | 0.69 | 0.213 | 1.69E-221 | chr12 |
| 437 | MESDC2 | 1.54E-225 | 0.44388467 | 0.442 | 0.001 | 4.52E-221 | chr15 |
| 616 | PTDSS1 | 2.65E-224 | 0.50650772 | 0.586 | 0.098 | 7.78E-220 | chr8 |
| 494 | NDUFA4L2 | 5.07E-224 | 1.68873016 | 0.522 | 0.063 | 1.49E-219 | chr12 |
| 770 | SYPL1 | 2.91E-223 | 0.55367418 | 0.643 | 0.147 | 8.54E-219 | chr7 |
| 536 | P4HB | 3.08E-223 | 0.84998202 | 0.819 | 0.443 | 9.04E-219 | chr17 |
| 270 | ENO1 | 3.16E-221 | 0.93666219 | 0.892 | 0.68 | 9.27E-217 | chr1 |
| 479 | MYL9 | 4.14E-221 | 0.75528127 | 0.54 | 0.059 | 1.21E-216 | chr20 |
| 255 | EGLN2 | 7.41E-221 | 0.46303749 | 0.474 | 0.021 | 2.17E-216 | chr19 |

| | | | | | | | |
|---|---|---|---|---|---|---|---|
| 754 | SQSTM1 | 9.45E-219 | 0.79747809 | 0.75 | 0.259 | 2.77E-214 | chr5 |
| 313 | GBAS | 9.52E-219 | 0.40265115 | 0.432 | 0.001 | 2.79E-214 | chr7 |
| 184 | COMMD6 | 1.53E-218 | 0.66542993 | 0.779 | 0.279 | 4.50E-214 | chr13 |
| 274 | EPB41L4A-AS1 | 1.83E-218 | 0.54105399 | 0.65 | 0.141 | 5.37E-214 | chr5 |
| 326 | GPAT2 | 2.44E-218 | 0.46078397 | 0.432 | 0.001 | 7.15E-214 | chr2 |
| 287 | FAM3C | 7.55E-218 | 0.53389597 | 0.636 | 0.137 | 2.22E-213 | chr7 |
| 135 | CD44 | 6.66E-216 | 0.54588626 | 0.635 | 0.129 | 1.95E-211 | chr11 |
| 100 | C19orf60 | 7.54E-215 | 0.39048194 | 0.426 | 0.001 | 2.21E-210 | chr19 |
| 303 | FMOD | 2.71E-213 | 0.64149165 | 0.638 | 0.148 | 7.97E-209 | chr1 |
| 329 | GRINA | 2.90E-213 | 0.58461304 | 0.668 | 0.178 | 8.51E-209 | chr8 |
| 863 | WHSC1L1 | 4.00E-213 | 0.3911322 | 0.423 | 0.001 | 1.17E-208 | chr8 |
| 567 | PKM | 1.02E-212 | 0.84448194 | 0.843 | 0.554 | 3.01E-208 | chr15 |
| 670 | RTFDC1 | 2.54E-212 | 0.36205014 | 0.422 | 0.001 | 7.46E-208 | chr20 |
| 214 | CYB5R3 | 4.47E-211 | 0.59090251 | 0.656 | 0.185 | 1.31E-206 | chr22 |
| 454 | MRPL37 | 1.97E-209 | 0.47976605 | 0.572 | 0.108 | 5.79E-205 | chr1 |
| 797 | TMEM261 | 1.78E-207 | 0.3695065 | 0.413 | 0 | 5.23E-203 | chr9 |
| 388 | KLF10 | 2.22E-207 | 0.67783367 | 0.621 | 0.152 | 6.50E-203 | chr8 |
| 875 | ZFAS1 | 2.71E-207 | 0.92724303 | 0.858 | 0.436 | 7.96E-203 | chr20 |
| 197 | CPSF3L | 1.25E-206 | 0.35256451 | 0.412 | 0 | 3.68E-202 | chr1 |
| 10 | ADSL | 3.71E-206 | 0.61837374 | 0.496 | 0.056 | 1.09E-201 | chr22 |
| 107 | C4orf3 | 4.47E-206 | 0.55035151 | 0.71 | 0.209 | 1.31E-201 | chr4 |
| 420 | LY6E | 1.27E-205 | 1.06104647 | 0.797 | 0.375 | 3.72E-201 | chr8 |
| 298 | FDX1 | 2.03E-204 | 0.46155668 | 0.502 | 0.057 | 5.96E-200 | chr11 |
| 290 | FAM92A1 | 2.63E-204 | 0.44623346 | 0.408 | 0 | 7.73E-200 | chr8 |
| 461 | MRPS6 | 2.99E-204 | 0.8688456 | 0.752 | 0.278 | 8.77E-200 | chr21 |
| 792 | TMEM173 | 7.53E-204 | 0.53514873 | 0.556 | 0.095 | 2.21E-199 | chr5 |
| 719 | SLC16A3 | 6.81E-203 | 0.61895549 | 0.558 | 0.094 | 2.00E-198 | chr17 |
| 640 | RCN1 | 1.47E-202 | 0.74817909 | 0.679 | 0.222 | 4.31E-198 | chr11 |
| 153 | CECR5 | 3.72E-201 | 0.35271437 | 0.403 | 0 | 1.09E-196 | chr22 |
| 103 | C1QTNF3 | 4.91E-201 | 0.63345583 | 0.528 | 0.075 | 1.44E-196 | chr5 |
| 643 | RGS3 | 7.99E-201 | 0.59363077 | 0.637 | 0.163 | 2.34E-196 | chr9 |
| 88 | C14orf1 | 1.58E-200 | 0.37301699 | 0.402 | 0 | 4.64E-196 | chr14 |
| 563 | PHPT1 | 3.43E-200 | 0.6473332 | 0.759 | 0.311 | 1.01E-195 | chr9 |
| 684 | SDC2 | 4.33E-200 | 0.72075902 | 0.718 | 0.275 | 1.27E-195 | chr8 |
| 327 | GPNMB | 3.33E-199 | 0.75364037 | 0.436 | 0.018 | 9.77E-195 | chr7 |
| 258 | EI24 | 1.65E-197 | 0.49016165 | 0.629 | 0.157 | 4.85E-193 | chr11 |
| 683 | SCPEP1 | 2.07E-197 | 0.43481408 | 0.545 | 0.091 | 6.08E-193 | chr17 |
| 210 | CTSL | 3.18E-197 | 0.50382456 | 0.538 | 0.085 | 9.33E-193 | chr9 |
| 92 | C16orf13 | 3.41E-197 | 0.33372098 | 0.397 | 0 | 1.00E-192 | chr16 |

| | | | | | | | |
|---|---|---|---|---|---|---|---|
| 408 | LINC00657 | 3.41E-197 | 0.33153085 | 0.397 | 0 | 1.00E-192 | chr20 |
| 522 | NUDT14 | 3.12E-196 | 0.50973343 | 0.553 | 0.107 | 9.16E-192 | chr14 |
| 406 | LINC00493 | 4.27E-196 | 0.32484625 | 0.397 | 0.001 | 1.25E-191 | chr20 |
| 309 | FXYD6 | 1.07E-195 | 0.4775172 | 0.553 | 0.102 | 3.15E-191 | chr11 |
| 826 | TUBB6 | 3.28E-195 | 0.51423635 | 0.566 | 0.113 | 9.63E-191 | chr18 |
| 85 | C11orf73 | 9.14E-195 | 0.32659491 | 0.394 | 0.001 | 2.68E-190 | chr11 |
| 62 | ATP5SL | 1.04E-194 | 0.313675 | 0.393 | 0 | 3.07E-190 | chr19 |
| 384 | KDELR2 | 1.50E-194 | 0.70225523 | 0.813 | 0.418 | 4.39E-190 | chr7 |
| 418 | LTBR | 1.74E-194 | 0.3624099 | 0.401 | 0.004 | 5.12E-190 | chr12 |
| 186 | COPRS | 3.20E-194 | 0.61574141 | 0.562 | 0.119 | 9.38E-190 | chr17 |
| 201 | CRISPLD1 | 9.62E-194 | 0.68224347 | 0.626 | 0.156 | 2.82E-189 | chr8 |
| 413 | LOXL1 | 1.90E-193 | 0.49814326 | 0.585 | 0.118 | 5.57E-189 | chr15 |
| 224 | DDIT4 | 1.51E-192 | 1.58347951 | 0.704 | 0.294 | 4.42E-188 | chr10 |
| 801 | TMEM55A | 2.25E-192 | 0.48109185 | 0.391 | 0.001 | 6.60E-188 | chr8 |
| 244 | DUSP2 | 5.97E-192 | 0.80077351 | 0.464 | 0.047 | 1.75E-187 | chr2 |
| 225 | DDT | 6.74E-192 | 0.70391519 | 0.691 | 0.245 | 1.98E-187 | chr22 |
| 685 | SDCBP | 6.02E-191 | 0.75007265 | 0.766 | 0.372 | 1.77E-186 | chr8 |
| 822 | TSPAN4 | 1.50E-190 | 0.63901847 | 0.584 | 0.136 | 4.41E-186 | chr11 |
| 208 | CTSD | 1.58E-190 | 0.59992396 | 0.673 | 0.209 | 4.64E-186 | chr11 |
| 187 | COPZ1 | 6.32E-190 | 0.44629682 | 0.623 | 0.157 | 1.85E-185 | chr12 |
| 561 | PHLDA1 | 1.64E-189 | 0.51964439 | 0.492 | 0.059 | 4.82E-185 | chr12 |
| 20 | ALDOA | 3.24E-189 | 0.93411885 | 0.642 | 0.209 | 9.52E-185 | chr16 |
| 64 | ATP6V0E1 | 4.18E-189 | 0.57219093 | 0.723 | 0.263 | 1.23E-184 | chr5 |
| 353 | HSPB1 | 4.48E-189 | 0.97199774 | 0.835 | 0.517 | 1.31E-184 | chr7 |
| 102 | C1orf43 | 6.50E-189 | 0.5074949 | 0.659 | 0.201 | 1.91E-184 | chr1 |
| 235 | DKK3 | 7.04E-189 | 0.46081228 | 0.431 | 0.026 | 2.07E-184 | chr11 |
| 838 | UQCR11 | 1.63E-188 | 0.6618102 | 0.796 | 0.402 | 4.79E-184 | chr19 |
| 386 | KIAA0101 | 8.37E-188 | 0.62043731 | 0.383 | 0.001 | 2.46E-183 | chr15 |
| 156 | CHCHD2 | 1.82E-187 | 0.61200529 | 0.896 | 0.68 | 5.33E-183 | chr7 |
| 207 | CTSB | 1.91E-187 | 0.66294768 | 0.647 | 0.193 | 5.60E-183 | chr8 |
| 648 | RHOC | 2.81E-187 | 0.60039312 | 0.711 | 0.26 | 8.24E-183 | chr1 |
| 596 | PPP2R4 | 2.45E-186 | 0.29172369 | 0.381 | 0.001 | 7.18E-182 | chr9 |
| 700 | SERPINE2 | 1.33E-185 | 0.90747631 | 0.638 | 0.178 | 3.89E-181 | chr2 |
| 841 | UQCRQ | 2.00E-185 | 0.63539226 | 0.787 | 0.387 | 5.87E-181 | chr5 |
| 70 | AXL | 2.14E-185 | 0.43170716 | 0.49 | 0.063 | 6.27E-181 | chr19 |
| 873 | ZFAND1 | 3.05E-185 | 0.36009825 | 0.524 | 0.086 | 8.93E-181 | chr8 |
| 179 | COL5A1 | 3.05E-185 | 0.50688523 | 0.626 | 0.15 | 8.94E-181 | chr9 |
| 155 | CFH | 8.98E-185 | 0.36042667 | 0.396 | 0.009 | 2.63E-180 | chr1 |
| 645 | RHBDD2 | 6.95E-184 | 0.40503892 | 0.541 | 0.103 | 2.04E-179 | chr7 |
| 421 | LY6K | 1.63E-183 | 0.4325352 | 0.378 | 0.002 | 4.77E-179 | chr8 |

| | | | | | | | |
|---|---|---|---|---|---|---|---|
| 491 | NDUFA11 | 3.65E-183 | 0.63021497 | 0.762 | 0.353 | 1.07E-178 | chr19 |
| 282 | FAM103A1 | 7.26E-183 | 0.27876336 | 0.374 | 0 | 2.13E-178 | chr15 |
| 595 | PPP2R1A | 2.97E-182 | 0.49937239 | 0.7 | 0.245 | 8.71E-178 | chr19 |
| 532 | OXA1L | 3.34E-182 | 0.41037816 | 0.595 | 0.139 | 9.79E-178 | chr14 |
| 409 | LITAF | 3.91E-182 | 0.56118718 | 0.718 | 0.248 | 1.15E-177 | chr16 |
| 94 | C17orf62 | 4.65E-182 | 0.30814401 | 0.372 | 0 | 1.36E-177 | chr17 |
| 477 | MYL12A | 7.22E-182 | 0.6320566 | 0.693 | 0.274 | 2.12E-177 | chr18 |
| 831 | TXNDC12 | 8.49E-182 | 0.46384279 | 0.603 | 0.153 | 2.49E-177 | chr1 |
| 397 | LAPTM4A | 8.88E-182 | 0.72723819 | 0.813 | 0.491 | 2.61E-177 | chr2 |
| 735 | SNAI2 | 2.05E-181 | 0.60929395 | 0.53 | 0.107 | 6.02E-177 | chr8 |
| 83 | BSG | 9.05E-181 | 0.72305135 | 0.756 | 0.352 | 2.66E-176 | chr19 |
| 281 | FAM101B | 1.19E-180 | 0.35840491 | 0.37 | 0 | 3.48E-176 | chr17 |
| 604 | PRR13 | 7.75E-180 | 0.42330211 | 0.58 | 0.137 | 2.27E-175 | chr12 |
| 283 | FAM127A | 1.20E-179 | 0.29318209 | 0.368 | 0 | 3.51E-175 | chrX |
| 525 | OAF | 1.86E-179 | 0.53766306 | 0.61 | 0.151 | 5.47E-175 | chr11 |
| 601 | PRKCDBP | 1.90E-179 | 0.45893556 | 0.368 | 0 | 5.56E-175 | chr11 |
| 415 | LSM1 | 2.18E-179 | 0.38801266 | 0.572 | 0.128 | 6.39E-175 | chr8 |
| 169 | CLEC2B | 2.36E-179 | 0.51429179 | 0.419 | 0.024 | 6.93E-175 | chr12 |
| 689 | SDHD | 3.37E-179 | 0.31548143 | 0.418 | 0.025 | 9.89E-175 | chr11 |
| 807 | TNFRSF1A | 3.98E-179 | 0.42454654 | 0.525 | 0.093 | 1.17E-174 | chr12 |
| 463 | MSC | 4.41E-179 | 0.61905904 | 0.386 | 0.011 | 1.29E-174 | chr8 |
| 233 | DGCR6L | 4.64E-179 | 0.41272021 | 0.539 | 0.107 | 1.36E-174 | chr22 |
| 369 | IL11RA | 5.59E-179 | 0.49319392 | 0.517 | 0.084 | 1.64E-174 | chr9 |
| 657 | RP11-386G11.10 | 5.59E-179 | 0.40532095 | 0.369 | 0.001 | 1.64E-174 | chr12 |
| 484 | NAB2 | 9.87E-179 | 0.35408277 | 0.446 | 0.042 | 2.89E-174 | chr12 |
| 65 | ATP6V1E1 | 1.04E-178 | 0.43680928 | 0.568 | 0.125 | 3.06E-174 | chr22 |
| 501 | NDUFC2 | 1.84E-178 | 0.63444414 | 0.746 | 0.334 | 5.41E-174 | chr11 |
| 696 | SEPN1 | 4.76E-178 | 0.27942505 | 0.366 | 0 | 1.40E-173 | chr1 |
| 555 | PGF | 9.77E-178 | 0.76096175 | 0.656 | 0.23 | 2.87E-173 | chr14 |
| 422 | LYPLA1 | 1.02E-177 | 0.45964743 | 0.593 | 0.151 | 3.00E-173 | chr8 |
| 302 | FKBP10 | 1.42E-177 | 0.56168569 | 0.647 | 0.197 | 4.16E-173 | chr17 |
| 16 | AK4 | 2.81E-177 | 0.34313267 | 0.434 | 0.036 | 8.24E-173 | chr1 |
| 796 | TMEM230 | 3.04E-177 | 0.42810754 | 0.643 | 0.183 | 8.92E-173 | chr20 |
| 549 | PEF1 | 4.17E-177 | 0.39997056 | 0.574 | 0.136 | 1.22E-172 | chr1 |
| 424 | MAF1 | 9.85E-177 | 0.48618182 | 0.686 | 0.232 | 2.89E-172 | chr8 |
| 679 | SCARB2 | 3.09E-176 | 0.37452038 | 0.5 | 0.075 | 9.05E-172 | chr4 |
| 448 | MMP24-AS1 | 3.38E-176 | 0.30203967 | 0.364 | 0.001 | 9.92E-172 | chr20 |
| 508 | NNAT | 4.85E-176 | 2.63589669 | 0.397 | 0.026 | 1.42E-171 | chr20 |
| 264 | ELN | 5.08E-176 | 0.66112461 | 0.551 | 0.116 | 1.49E-171 | chr7 |

| | | | | | | | |
|---|---|---|---|---|---|---|---|
| 788 | TMEM109 | 1.30E-175 | 0.41507345 | 0.568 | 0.133 | 3.81E-171 | chr11 |
| 653 | RNF212 | 2.87E-175 | 0.34836221 | 0.382 | 0.009 | 8.42E-171 | chr4 |
| 173 | CNIH1 | 3.55E-175 | 0.39575641 | 0.619 | 0.159 | 1.04E-170 | chr14 |
| 757 | SSNA1 | 6.72E-175 | 0.47501144 | 0.639 | 0.196 | 1.97E-170 | chr9 |
| 531 | OSTC | 1.08E-174 | 0.57549971 | 0.759 | 0.317 | 3.18E-170 | chr4 |
| 385 | KDELR3 | 1.19E-174 | 0.41206814 | 0.493 | 0.079 | 3.50E-170 | chr22 |
| 857 | WBSCR16 | 1.83E-174 | 0.25340275 | 0.361 | 0.001 | 5.37E-170 | chr7 |
| 372 | IMPAD1 | 1.93E-174 | 0.47491121 | 0.571 | 0.14 | 5.67E-170 | chr8 |
| 110 | C7orf49 | 4.52E-174 | 0.27652564 | 0.359 | 0 | 1.32E-169 | chr7 |
| 802 | TMEM70 | 8.82E-173 | 0.46401582 | 0.591 | 0.155 | 2.59E-168 | chr8 |
| 13 | AGPAT2 | 2.32E-172 | 0.40271109 | 0.603 | 0.156 | 6.79E-168 | chr9 |
| 228 | DDX54 | 1.63E-171 | 0.32699702 | 0.508 | 0.086 | 4.79E-167 | chr12 |
| 658 | RP11-395G23.3 | 1.67E-171 | 0.33956679 | 0.355 | 0 | 4.89E-167 | chr8 |
| 544 | PDLIM3 | 7.08E-171 | 0.56236994 | 0.383 | 0.017 | 2.08E-166 | chr4 |
| 15 | AK1 | 1.79E-170 | 0.40853922 | 0.494 | 0.084 | 5.26E-166 | chr9 |
| 9 | ADPRHL2 | 3.15E-170 | 0.33729624 | 0.514 | 0.091 | 9.23E-166 | chr1 |
| 192 | COX7A2L | 3.74E-170 | 0.57097475 | 0.744 | 0.337 | 1.10E-165 | chr2 |
| 694 | SELT | 7.27E-170 | 0.25456323 | 0.353 | 0.001 | 2.13E-165 | chr3 |
| 756 | SRSF9 | 8.11E-170 | 0.59605916 | 0.763 | 0.372 | 2.38E-165 | chr12 |
| 147 | CDK4 | 9.04E-170 | 0.52931938 | 0.686 | 0.26 | 2.65E-165 | chr12 |
| 157 | CHID1 | 9.76E-170 | 0.38844424 | 0.573 | 0.135 | 2.86E-165 | chr11 |
| 540 | PARVB | 1.11E-169 | 0.33059496 | 0.46 | 0.059 | 3.27E-165 | chr22 |
| 130 | CCND3 | 1.38E-169 | 0.39570998 | 0.514 | 0.097 | 4.06E-165 | chr6 |
| 139 | CD82 | 6.61E-169 | 0.33409535 | 0.392 | 0.022 | 1.94E-164 | chr11 |
| 748 | SPG21 | 1.30E-168 | 0.36790145 | 0.542 | 0.113 | 3.82E-164 | chr15 |
| 140 | CD99 | 2.36E-168 | 0.75489411 | 0.753 | 0.31 | 6.92E-164 | chrX |
| 288 | FAM46A | 2.66E-168 | 0.34161612 | 0.351 | 0.001 | 7.79E-164 | chr6 |
| 3 | ABCF2 | 5.83E-168 | 0.27897416 | 0.371 | 0.01 | 1.71E-163 | chr7 |
| 304 | FNTA | 1.33E-167 | 0.35665533 | 0.561 | 0.127 | 3.89E-163 | chr8 |
| 675 | S100A2 | 1.46E-166 | 0.73238921 | 0.364 | 0.009 | 4.29E-162 | chr1 |
| 432 | MDH2 | 2.45E-166 | 0.5273288 | 0.736 | 0.313 | 7.20E-162 | chr7 |
| 872 | ZDHHC4 | 6.31E-166 | 0.36684386 | 0.538 | 0.113 | 1.85E-161 | chr7 |
| 149 | CDKN1A | 7.02E-166 | 0.49984332 | 0.536 | 0.114 | 2.06E-161 | chr6 |
| 504 | NEK6 | 1.39E-165 | 0.32402013 | 0.466 | 0.067 | 4.09E-161 | chr9 |
| 151 | CDKN2C | 1.85E-165 | 0.43758845 | 0.494 | 0.09 | 5.43E-161 | chr1 |
| 185 | COMT | 3.95E-165 | 0.2922044 | 0.462 | 0.063 | 1.16E-160 | chr22 |
| 82 | BRK1 | 4.84E-164 | 0.82556526 | 0.731 | 0.356 | 1.42E-159 | chr3 |
| 605 | PRR5 | 5.17E-164 | 0.35712212 | 0.449 | 0.06 | 1.52E-159 | chr22 |
| 528 | OLFML3 | 6.57E-164 | 0.44737404 | 0.514 | 0.101 | 1.93E-159 | chr1 |
| 762 | ST3GAL4 | 8.89E-164 | 0.31467404 | 0.394 | 0.027 | 2.61E-159 | chr11 |

| | | | | | | | |
|---|---|---|---|---|---|---|---|
| 279 | FABP3 | 1.58E-163 | 0.44074999 | 0.369 | 0.013 | 4.63E-159 | chr1 |
| 830 | TXN2 | 1.60E-163 | 0.43328547 | 0.654 | 0.212 | 4.70E-159 | chr22 |
| 46 | ASPH | 2.56E-163 | 0.46193544 | 0.621 | 0.176 | 7.51E-159 | chr8 |
| 8 | ADM | 3.01E-163 | 0.68717324 | 0.556 | 0.133 | 8.84E-159 | chr11 |
| 712 | SHISA5 | 4.30E-163 | 0.42832808 | 0.594 | 0.165 | 1.26E-158 | chr3 |
| 781 | TMBIM6 | 8.56E-163 | 0.56506751 | 0.776 | 0.418 | 2.51E-158 | chr12 |
| 603 | PRNP | 2.87E-162 | 0.39696952 | 0.465 | 0.071 | 8.41E-158 | chr20 |
| 152 | CEBPD | 3.05E-162 | 1.06027707 | 0.648 | 0.252 | 8.94E-158 | chr8 |
| 747 | SPG20 | 3.70E-162 | 0.25405838 | 0.34 | 0.001 | 1.09E-157 | chr13 |
| 829 | TXN | 8.03E-162 | 0.58501208 | 0.776 | 0.401 | 2.36E-157 | chr9 |
| 798 | TMEM43 | 8.09E-162 | 0.31761449 | 0.445 | 0.057 | 2.38E-157 | chr3 |
| 780 | TM2D1 | 8.31E-162 | 0.3531989 | 0.576 | 0.138 | 2.44E-157 | chr1 |
| 275 | EPHX1 | 9.26E-162 | 0.39114301 | 0.448 | 0.06 | 2.72E-157 | chr1 |
| 587 | POP4 | 1.34E-161 | 0.26000377 | 0.411 | 0.037 | 3.94E-157 | chr19 |
| 30 | ANXA5 | 3.91E-161 | 0.67529011 | 0.734 | 0.297 | 1.15E-156 | chr4 |
| 868 | YARS | 4.24E-161 | 0.38140493 | 0.51 | 0.1 | 1.25E-156 | chr1 |
| 624 | PVRL2 | 5.47E-161 | 0.26184762 | 0.337 | 0 | 1.61E-156 | chr19 |
| 768 | SUN1 | 6.19E-161 | 0.53603043 | 0.507 | 0.095 | 1.82E-156 | chr7 |
| 818 | TRIOBP | 9.01E-161 | 0.34116653 | 0.484 | 0.083 | 2.64E-156 | chr22 |
| 419 | LUM | 9.33E-161 | 1.11413273 | 0.436 | 0.062 | 2.74E-156 | chr12 |
| 717 | SIX1 | 1.16E-160 | 0.73355834 | 0.623 | 0.22 | 3.40E-156 | chr14 |
| 803 | TMEM9 | 1.92E-160 | 0.44962876 | 0.574 | 0.156 | 5.62E-156 | chr1 |
| 449 | MOXD1 | 3.31E-160 | 0.37060589 | 0.429 | 0.052 | 9.72E-156 | chr6 |
| 315 | GGH | 5.00E-160 | 0.69694807 | 0.566 | 0.175 | 1.47E-155 | chr8 |
| 115 | C9orf142 | 1.89E-159 | 0.26582085 | 0.334 | 0 | 5.54E-155 | chr9 |
| 564 | PIH1D1 | 4.04E-159 | 0.29358501 | 0.426 | 0.045 | 1.19E-154 | chr19 |
| 621 | PTPMT1 | 6.24E-159 | 0.37568489 | 0.506 | 0.105 | 1.83E-154 | chr11 |
| 305 | FRZB | 8.20E-159 | 0.45365551 | 0.352 | 0.009 | 2.41E-154 | chr2 |
| 816 | TRIAP1 | 1.08E-158 | 0.33121157 | 0.507 | 0.099 | 3.16E-154 | chr12 |
| 697 | SEPP1 | 1.10E-158 | 0.65140948 | 0.333 | 0 | 3.24E-154 | chr5 |
| 859 | WDR13 | 1.25E-158 | 0.31214642 | 0.477 | 0.079 | 3.66E-154 | chrX |
| 40 | ARL5A | 1.35E-158 | 0.34058619 | 0.528 | 0.116 | 3.96E-154 | chr2 |
| 174 | CNN3 | 2.03E-158 | 0.70198483 | 0.74 | 0.355 | 5.96E-154 | chr1 |
| 4 | ACADVL | 4.25E-158 | 0.43509164 | 0.574 | 0.144 | 1.25E-153 | chr17 |
| 443 | MINOS1 | 1.14E-157 | 0.66443365 | 0.618 | 0.24 | 3.34E-153 | chr1 |
| 194 | CPE | 1.90E-157 | 0.72651591 | 0.69 | 0.261 | 5.58E-153 | chr4 |
| 455 | MRPL41 | 5.54E-157 | 0.40619762 | 0.607 | 0.189 | 1.63E-152 | chr9 |
| 440 | MFGE8 | 6.36E-157 | 0.4377188 | 0.553 | 0.137 | 1.87E-152 | chr15 |
| 680 | SCARF2 | 1.01E-156 | 0.32944038 | 0.402 | 0.038 | 2.95E-152 | chr22 |
| 219 | DARS | 1.42E-156 | 0.36643336 | 0.595 | 0.157 | 4.15E-152 | chr2 |

| | | | | | | | |
|---|---|---|---|---|---|---|---|
| 552 | PEX2 | 1.63E-156 | 0.35038597 | 0.574 | 0.144 | 4.79E-152 | chr8 |
| 109 | C6orf48 | 1.82E-156 | 0.74886502 | 0.674 | 0.305 | 5.35E-152 | chr6 |
| 112 | C7orf55 | 3.10E-156 | 0.27637381 | 0.33 | 0.001 | 9.11E-152 | chr7 |
| 213 | CYB5R1 | 3.23E-156 | 0.38048344 | 0.417 | 0.048 | 9.48E-152 | chr1 |
| 785 | TMED3 | 3.79E-156 | 0.38913049 | 0.648 | 0.205 | 1.11E-151 | chr15 |
| 705 | SERTAD3 | 6.33E-156 | 0.30534648 | 0.46 | 0.072 | 1.86E-151 | chr19 |
| 759 | SSR2 | 6.60E-156 | 0.64741441 | 0.815 | 0.43 | 1.94E-151 | chr1 |
| 647 | RHOBTB3 | 9.93E-156 | 0.54083269 | 0.657 | 0.248 | 2.91E-151 | chr5 |
| 771 | TAF1D | 1.04E-155 | 0.4543275 | 0.637 | 0.195 | 3.04E-151 | chr11 |
| 867 | XRCC6 | 1.18E-155 | 0.60659257 | 0.735 | 0.32 | 3.46E-151 | chr22 |
| 876 | ZFYVE21 | 2.11E-155 | 0.29214716 | 0.486 | 0.087 | 6.20E-151 | chr14 |
| 101 | C19orf70 | 2.30E-155 | 0.40318035 | 0.582 | 0.167 | 6.74E-151 | chr19 |
| 23 | ANAPC16 | 2.74E-155 | 0.44437519 | 0.653 | 0.224 | 8.03E-151 | chr10 |
| 394 | LAMP1 | 3.47E-155 | 0.46866837 | 0.662 | 0.234 | 1.02E-150 | chr13 |
| 509 | NNMT | 3.99E-155 | 0.58480447 | 0.372 | 0.022 | 1.17E-150 | chr11 |
| 183 | COMMD5 | 6.04E-155 | 0.32328746 | 0.5 | 0.096 | 1.77E-150 | chr8 |
| 652 | RNF187 | 7.34E-155 | 0.42211331 | 0.63 | 0.191 | 2.15E-150 | chr1 |
| 594 | PPP1R15A | 8.90E-155 | 0.73260616 | 0.675 | 0.26 | 2.61E-150 | chr19 |
| 148 | CDK6 | 9.71E-154 | 0.37902021 | 0.53 | 0.122 | 2.85E-149 | chr7 |
| 426 | MAGED2 | 2.09E-153 | 0.95374726 | 0.735 | 0.375 | 6.13E-149 | chrX |
| 25 | ANGPTL4 | 7.75E-153 | 0.76648693 | 0.523 | 0.119 | 2.27E-148 | chr19 |
| 296 | FBXW5 | 1.34E-152 | 0.33190269 | 0.543 | 0.133 | 3.93E-148 | chr9 |
| 215 | CYBRD1 | 1.43E-152 | 0.28151924 | 0.391 | 0.036 | 4.19E-148 | chr2 |
| 545 | PDLIM4 | 1.57E-152 | 0.47054457 | 0.627 | 0.202 | 4.59E-148 | chr5 |
| 745 | SPATC1L | 3.65E-152 | 0.32722322 | 0.442 | 0.066 | 1.07E-147 | chr21 |
| 79 | BOC | 3.82E-152 | 0.45486888 | 0.537 | 0.132 | 1.12E-147 | chr3 |
| 150 | CDKN2A | 3.88E-152 | 0.65235759 | 0.325 | 0.002 | 1.14E-147 | chr9 |
| 444 | MKKS | 7.01E-152 | 0.27993622 | 0.452 | 0.071 | 2.06E-147 | chr20 |
| 535 | P4HA2 | 1.13E-151 | 0.37684844 | 0.557 | 0.144 | 3.33E-147 | chr5 |
| 614 | PSMG2 | 1.24E-151 | 0.32445638 | 0.527 | 0.118 | 3.65E-147 | chr18 |
| 163 | CISD2 | 1.39E-151 | 0.30431345 | 0.527 | 0.118 | 4.07E-147 | chr4 |
| 460 | MRPS28 | 1.55E-151 | 0.28120423 | 0.448 | 0.067 | 4.55E-147 | chr8 |
| 618 | PTGES2 | 3.57E-151 | 0.31212296 | 0.472 | 0.089 | 1.05E-146 | chr9 |
| 257 | EHD2 | 4.73E-151 | 0.32728586 | 0.435 | 0.06 | 1.39E-146 | chr19 |
| 805 | TMX2 | 8.03E-151 | 0.3086041 | 0.479 | 0.093 | 2.36E-146 | chr11 |
| 263 | EIF5A | 8.05E-151 | 0.61862458 | 0.733 | 0.388 | 2.36E-146 | chr17 |
| 231 | DES | 1.03E-150 | 0.91698014 | 0.321 | 0.001 | 3.03E-146 | chr2 |
| 182 | COL6A3 | 1.53E-150 | 0.70763877 | 0.585 | 0.164 | 4.50E-146 | chr2 |
| 430 | MCOLN3 | 3.14E-150 | 0.39719486 | 0.393 | 0.041 | 9.22E-146 | chr1 |
| 204 | CSTB | 3.57E-150 | 0.48688221 | 0.724 | 0.278 | 1.05E-145 | chr21 |

| | | | | | | | |
|---|---|---|---|---|---|---|---|
| 355 | HSPG2 | 4.32E-150 | 0.45811381 | 0.49 | 0.099 | 1.27E-145 | chr1 |
| 2 | A2M | 5.12E-150 | 0.45981298 | 0.366 | 0.022 | 1.50E-145 | chr12 |
| 29 | ANXA2 | 5.57E-150 | 0.85719364 | 0.816 | 0.495 | 1.63E-145 | chr15 |
| 626 | PXMP2 | 5.63E-150 | 0.32367117 | 0.477 | 0.089 | 1.65E-145 | chr12 |
| 354 | HSPB6 | 5.83E-150 | 0.25448382 | 0.323 | 0.002 | 1.71E-145 | chr19 |
| 189 | COX6A1 | 8.91E-150 | 0.53091851 | 0.8 | 0.523 | 2.61E-145 | chr12 |
| 617 | PTGDS | 1.16E-149 | 0.90626002 | 0.33 | 0.007 | 3.41E-145 | chr9 |
| 773 | TATDN1 | 2.33E-149 | 0.39792519 | 0.652 | 0.23 | 6.83E-145 | chr8 |
| 469 | MT1X | 3.76E-149 | 0.81305954 | 0.691 | 0.267 | 1.10E-144 | chr16 |
| 392 | KXD1 | 4.84E-149 | 0.32666606 | 0.57 | 0.151 | 1.42E-144 | chr19 |
| 230 | DEGS1 | 1.13E-148 | 0.35870189 | 0.459 | 0.08 | 3.30E-144 | chr1 |
| 810 | TOMM22 | 1.25E-148 | 0.44920423 | 0.606 | 0.199 | 3.67E-144 | chr22 |
| 250 | ECHS1 | 1.47E-148 | 0.5494558 | 0.658 | 0.254 | 4.31E-144 | chr10 |
| 882 | ZYX | 1.69E-148 | 0.4236489 | 0.57 | 0.169 | 4.97E-144 | chr7 |
| 848 | VAMP3 | 1.88E-148 | 0.25702943 | 0.417 | 0.052 | 5.52E-144 | chr1 |
| 438 | MFAP2 | 2.09E-148 | 0.55773505 | 0.746 | 0.328 | 6.12E-144 | chr1 |
| 452 | MRPL13 | 2.13E-148 | 0.44241323 | 0.664 | 0.25 | 6.26E-144 | chr8 |
| 108 | C5orf15 | 2.47E-148 | 0.27211327 | 0.398 | 0.043 | 7.24E-144 | chr5 |
| 790 | TMEM123 | 3.00E-148 | 0.33677557 | 0.422 | 0.058 | 8.81E-144 | chr11 |
| 436 | MED30 | 4.79E-148 | 0.38954932 | 0.512 | 0.12 | 1.41E-143 | chr8 |
| 325 | GPAA1 | 5.37E-148 | 0.42437447 | 0.672 | 0.234 | 1.57E-143 | chr8 |
| 842 | URM1 | 6.36E-148 | 0.28038316 | 0.503 | 0.107 | 1.87E-143 | chr9 |
| 247 | EBAG9 | 9.19E-148 | 0.30161485 | 0.531 | 0.122 | 2.70E-143 | chr8 |
| 512 | NPM3 | 1.83E-147 | 0.35381069 | 0.545 | 0.144 | 5.36E-143 | chr10 |
| 524 | NXPH4 | 1.93E-147 | 0.41927836 | 0.375 | 0.033 | 5.66E-143 | chr12 |
| 472 | MTHFD2 | 2.28E-147 | 0.43631277 | 0.539 | 0.139 | 6.69E-143 | chr2 |
| 97 | C19orf12 | 5.95E-147 | 0.25211965 | 0.372 | 0.029 | 1.75E-142 | chr19 |
| 741 | SNRPN | 6.77E-147 | 0.60035808 | 0.592 | 0.207 | 1.98E-142 | chr15 |
| 598 | PRAF2 | 7.68E-147 | 0.30275983 | 0.462 | 0.084 | 2.25E-142 | chrX |
| 795 | TMEM222 | 9.79E-147 | 0.29368742 | 0.477 | 0.092 | 2.87E-142 | chr1 |
| 599 | PRAME | 2.50E-146 | 0.59638411 | 0.576 | 0.188 | 7.32E-142 | chr22 |
| 660 | RPA3 | 3.93E-146 | 0.3883885 | 0.568 | 0.16 | 1.15E-141 | chr7 |
| 374 | ISLR | 7.19E-146 | 0.43770006 | 0.581 | 0.165 | 2.11E-141 | chr15 |
| 162 | CISD1 | 8.59E-146 | 0.32451863 | 0.484 | 0.099 | 2.52E-141 | chr10 |
| 828 | TUG1 | 1.25E-145 | 0.29008292 | 0.337 | 0.011 | 3.68E-141 | chr22 |
| 569 | PLAGL1 | 1.34E-145 | 0.33714539 | 0.469 | 0.09 | 3.92E-141 | chr6 |
| 456 | MRPL55 | 1.48E-145 | 0.36350998 | 0.606 | 0.184 | 4.33E-141 | chr1 |
| 655 | RNMT | 4.82E-145 | 0.51720943 | 0.607 | 0.183 | 1.41E-140 | chr18 |
| 736 | SNAPIN | 7.88E-145 | 0.30765684 | 0.484 | 0.098 | 2.31E-140 | chr1 |
| 28 | ANXA11 | 1.07E-144 | 0.32874237 | 0.539 | 0.14 | 3.15E-140 | chr10 |

| | | | | | | | |
|---|---|---|---|---|---|---|---|
| 586 | PON2 | 2.64E-144 | 0.26980173 | 0.424 | 0.061 | 7.76E-140 | chr7 |
| 764 | STOM | 4.05E-144 | 0.26970604 | 0.427 | 0.062 | 1.19E-139 | chr9 |
| 649 | RNASEH2A | 9.69E-144 | 0.27155968 | 0.373 | 0.034 | 2.84E-139 | chr19 |
| 41 | ARL6IP4 | 1.36E-143 | 0.57177384 | 0.768 | 0.463 | 3.99E-139 | chr12 |
| 470 | MTFP1 | 2.33E-143 | 0.27527746 | 0.331 | 0.011 | 6.83E-139 | chr22 |
| 732 | SMIM3 | 3.72E-143 | 0.35482817 | 0.389 | 0.044 | 1.09E-138 | chr5 |
| 855 | VOPP1 | 4.08E-143 | 0.34064707 | 0.549 | 0.141 | 1.20E-138 | chr7 |
| 427 | MAN1B1 | 1.39E-142 | 0.28171186 | 0.434 | 0.067 | 4.08E-138 | chr9 |
| 74 | BCAP31 | 1.52E-142 | 0.38676314 | 0.653 | 0.222 | 4.46E-138 | chrX |
| 165 | CITED4 | 1.76E-142 | 0.42723461 | 0.439 | 0.075 | 5.16E-138 | chr1 |
| 330 | GRN | 2.55E-142 | 0.60628832 | 0.649 | 0.248 | 7.49E-138 | chr17 |
| 557 | PGM1 | 5.14E-142 | 0.3035614 | 0.507 | 0.116 | 1.51E-137 | chr1 |
| 170 | CLNS1A | 6.83E-142 | 0.38454022 | 0.633 | 0.22 | 2.01E-137 | chr11 |
| 416 | LSM10 | 3.44E-141 | 0.31394037 | 0.537 | 0.143 | 1.01E-136 | chr1 |
| 733 | SMIM7 | 5.23E-141 | 0.38031054 | 0.614 | 0.194 | 1.53E-136 | chr19 |
| 492 | NDUFA13 | 5.54E-141 | 0.54603226 | 0.769 | 0.417 | 1.63E-136 | chr19 |
| 24 | ANGPTL2 | 7.80E-141 | 0.35992391 | 0.455 | 0.083 | 2.29E-136 | chr9 |
| 678 | SAT2 | 9.89E-141 | 0.45217208 | 0.682 | 0.252 | 2.90E-136 | chr17 |
| 442 | MGP | 1.02E-140 | 1.21926532 | 0.789 | 0.361 | 2.99E-136 | chr12 |
| 630 | RAB34 | 1.07E-140 | 0.4592966 | 0.735 | 0.325 | 3.15E-136 | chr17 |
| 363 | IFI6 | 1.26E-140 | 0.40250312 | 0.409 | 0.06 | 3.70E-136 | chr1 |
| 480 | MYLPF | 2.70E-140 | 1.22964522 | 0.329 | 0.016 | 7.93E-136 | chr16 |
| 154 | CERCAM | 3.60E-140 | 0.26125473 | 0.396 | 0.05 | 1.06E-135 | chr9 |
| 239 | DPM2 | 4.21E-140 | 0.27706269 | 0.497 | 0.109 | 1.24E-135 | chr9 |
| 578 | PLXNB2 | 8.11E-140 | 0.35211788 | 0.488 | 0.105 | 2.38E-135 | chr22 |
| 126 | CCDC8 | 1.19E-139 | 0.26826914 | 0.445 | 0.078 | 3.50E-135 | chr19 |
| 488 | NDN | 1.89E-139 | 0.56636558 | 0.654 | 0.266 | 5.53E-135 | chr15 |
| 119 | CAPN2 | 3.67E-139 | 0.318975 | 0.515 | 0.123 | 1.08E-134 | chr1 |
| 767 | SULF1 | 1.03E-138 | 0.34704569 | 0.376 | 0.039 | 3.03E-134 | chr8 |
| 534 | P4HA1 | 1.41E-138 | 0.39828637 | 0.626 | 0.196 | 4.14E-134 | chr10 |
| 743 | SPAG7 | 1.50E-138 | 0.28309254 | 0.599 | 0.181 | 4.39E-134 | chr17 |
| 362 | IFI16 | 2.71E-138 | 0.45482256 | 0.43 | 0.074 | 7.96E-134 | chr1 |
| 393 | LACTB2 | 3.35E-138 | 0.3359975 | 0.441 | 0.081 | 9.82E-134 | chr8 |
| 682 | SCOC | 7.61E-138 | 0.26339295 | 0.538 | 0.14 | 2.23E-133 | chr4 |
| 704 | SERTAD1 | 1.88E-137 | 0.42594768 | 0.595 | 0.188 | 5.53E-133 | chr19 |
| 121 | CAV1 | 1.95E-137 | 0.40530892 | 0.478 | 0.093 | 5.73E-133 | chr7 |
| 405 | LINC00152 | 2.35E-137 | 0.37465756 | 0.295 | 0 | 6.90E-133 | chr2 |
| 851 | VDAC1 | 2.53E-137 | 0.48092979 | 0.709 | 0.316 | 7.42E-133 | chr5 |
| 118 | CAMLG | 2.64E-137 | 0.35310932 | 0.617 | 0.194 | 7.76E-133 | chr5 |
| 677 | SAMM50 | 4.53E-137 | 0.32908873 | 0.531 | 0.145 | 1.33E-132 | chr22 |

| | | | | | | | |
|---|---|---|---|---|---|---|---|
| 159 | CHRNA1 | 8.23E-137 | 0.62827066 | 0.316 | 0.012 | 2.42E-132 | chr2 |
| 529 | OS9 | 2.76E-136 | 0.38521044 | 0.631 | 0.218 | 8.10E-132 | chr12 |
| 99 | C19orf53 | 2.86E-136 | 0.43358277 | 0.763 | 0.356 | 8.40E-132 | chr19 |
| 339 | HILPDA | 3.23E-136 | 0.4578298 | 0.532 | 0.138 | 9.49E-132 | chr7 |
| 708 | SH3GL1 | 5.26E-136 | 0.33086085 | 0.542 | 0.148 | 1.54E-131 | chr19 |
| 212 | CXXC5 | 6.28E-136 | 0.39613524 | 0.584 | 0.18 | 1.84E-131 | chr5 |
| 686 | SDF2L1 | 1.12E-135 | 0.44331254 | 0.579 | 0.194 | 3.29E-131 | chr22 |
| 217 | CYSTM1 | 1.13E-135 | 0.31843308 | 0.423 | 0.071 | 3.30E-131 | chr5 |
| 627 | PYURF | 1.19E-135 | 0.31031187 | 0.576 | 0.168 | 3.50E-131 | chr4 |
| 663 | RPS19BP1 | 1.31E-135 | 0.43066613 | 0.707 | 0.303 | 3.83E-131 | chr22 |
| 203 | CRTAP | 1.35E-135 | 0.3571216 | 0.591 | 0.183 | 3.97E-131 | chr3 |
| 814 | TPM2 | 3.40E-135 | 0.58307698 | 0.771 | 0.417 | 9.98E-131 | chr9 |
| 457 | MRPS12 | 3.49E-135 | 0.30214162 | 0.57 | 0.169 | 1.02E-130 | chr19 |
| 450 | MPG | 4.18E-135 | 0.32561268 | 0.551 | 0.157 | 1.23E-130 | chr16 |
| 502 | NDUFS5 | 4.96E-135 | 0.62061342 | 0.774 | 0.43 | 1.46E-130 | chr1 |
| 336 | HEXA | 7.52E-135 | 0.30995123 | 0.538 | 0.145 | 2.21E-130 | chr15 |
| 778 | THY1 | 1.30E-134 | 0.518432 | 0.475 | 0.118 | 3.83E-130 | chr11 |
| 114 | C8orf33 | 1.91E-134 | 0.38861495 | 0.68 | 0.239 | 5.61E-130 | chr8 |
| 687 | SDHAF2 | 3.11E-134 | 0.29950341 | 0.477 | 0.106 | 9.12E-130 | chr11 |
| 481 | MYOD1 | 4.93E-134 | 0.55675501 | 0.296 | 0.003 | 1.45E-129 | chr11 |
| 370 | IL32 | 5.13E-134 | 0.48741346 | 0.315 | 0.013 | 1.51E-129 | chr16 |
| 709 | SHC1 | 5.32E-134 | 0.35575513 | 0.561 | 0.168 | 1.56E-129 | chr1 |
| 441 | MGMT | 5.71E-134 | 0.30359788 | 0.555 | 0.155 | 1.68E-129 | chr10 |
| 295 | FBXO7 | 7.57E-134 | 0.33037822 | 0.523 | 0.14 | 2.22E-129 | chr22 |
| 471 | MTFR1L | 1.57E-133 | 0.30082807 | 0.503 | 0.127 | 4.61E-129 | chr1 |
| 597 | PPT1 | 2.04E-133 | 0.38963394 | 0.593 | 0.203 | 5.99E-129 | chr1 |
| 793 | TMEM176B | 2.13E-133 | 0.41928829 | 0.373 | 0.047 | 6.26E-129 | chr7 |
| 548 | PEBP1 | 2.19E-133 | 0.55443232 | 0.744 | 0.359 | 6.44E-129 | chr12 |
| 706 | SGCA | 2.80E-133 | 0.61426174 | 0.29 | 0.001 | 8.20E-129 | chr17 |
| 750 | SPOCK2 | 7.81E-133 | 0.73148223 | 0.32 | 0.018 | 2.29E-128 | chr10 |
| 134 | CD320 | 7.83E-133 | 0.3527842 | 0.573 | 0.176 | 2.30E-128 | chr19 |
| 879 | ZNF511 | 9.32E-133 | 0.31232751 | 0.505 | 0.129 | 2.73E-128 | chr10 |
| 667 | RRS1 | 1.78E-132 | 0.25637243 | 0.455 | 0.091 | 5.22E-128 | chr8 |
| 345 | HLA-E | 4.16E-132 | 0.54144809 | 0.657 | 0.252 | 1.22E-127 | chr6 |
| 740 | SNRNP40 | 4.53E-132 | 0.25552191 | 0.514 | 0.133 | 1.33E-127 | chr1 |
| 755 | SRM | 4.79E-132 | 0.51434452 | 0.704 | 0.317 | 1.40E-127 | chr1 |
| 106 | C22orf39 | 5.88E-132 | 0.25892496 | 0.429 | 0.076 | 1.73E-127 | chr22 |
| 338 | HIGD2A | 2.62E-131 | 0.3456358 | 0.627 | 0.215 | 7.68E-127 | chr5 |
| 702 | SERPINH1 | 3.63E-131 | 0.61009802 | 0.777 | 0.39 | 1.07E-126 | chr11 |
| 556 | PGK1 | 5.49E-131 | 0.53941441 | 0.804 | 0.462 | 1.61E-126 | chrX |

| | | | | | | | |
|---|---|---|---|---|---|---|---|
| 131 | CCNI | 7.15E-131 | 0.52354931 | 0.804 | 0.475 | 2.10E-126 | chr4 |
| 291 | FASTK | 1.03E-130 | 0.32383058 | 0.572 | 0.179 | 3.03E-126 | chr7 |
| 482 | MZT2A | 1.18E-130 | 0.42727375 | 0.749 | 0.336 | 3.46E-126 | chr2 |
| 410 | LMCD1 | 1.53E-130 | 0.31160448 | 0.419 | 0.074 | 4.49E-126 | chr3 |
| 133 | CD164 | 1.60E-130 | 0.39019627 | 0.677 | 0.274 | 4.69E-126 | chr6 |
| 350 | HNRNPA1 | 2.05E-130 | 0.54121878 | 0.866 | 0.645 | 6.00E-126 | chr12 |
| 371 | IMPA2 | 2.21E-130 | 0.36114514 | 0.405 | 0.07 | 6.48E-126 | chr18 |
| 497 | NDUFB2 | 2.40E-130 | 0.51419285 | 0.765 | 0.394 | 7.04E-126 | chr7 |
| 789 | TMEM119 | 4.97E-130 | 0.2523167 | 0.331 | 0.026 | 1.46E-125 | chr12 |
| 846 | UTP23 | 5.57E-130 | 0.32496031 | 0.563 | 0.163 | 1.63E-125 | chr8 |
| 226 | DDX18 | 7.55E-130 | 0.38232414 | 0.705 | 0.262 | 2.21E-125 | chr2 |
| 716 | SIVA1 | 1.31E-129 | 0.46135681 | 0.742 | 0.341 | 3.84E-125 | chr14 |
| 265 | ELP5 | 1.60E-129 | 0.28167948 | 0.423 | 0.077 | 4.70E-125 | chr17 |
| 429 | MARVELD1 | 1.74E-129 | 0.3134841 | 0.512 | 0.131 | 5.11E-125 | chr10 |
| 777 | TGIF1 | 2.29E-129 | 0.50728308 | 0.48 | 0.13 | 6.72E-125 | chr18 |
| 301 | FHL2 | 2.55E-129 | 0.48761389 | 0.413 | 0.078 | 7.49E-125 | chr2 |
| 794 | TMEM203 | 5.50E-129 | 0.25666202 | 0.545 | 0.15 | 1.61E-124 | chr9 |
| 18 | AKR1C3 | 5.86E-129 | 0.30403611 | 0.38 | 0.054 | 1.72E-124 | chr10 |
| 266 | EMC10 | 1.20E-128 | 0.33815281 | 0.619 | 0.209 | 3.51E-124 | chr19 |
| 715 | SIRT2 | 3.24E-128 | 0.28722035 | 0.464 | 0.106 | 9.51E-124 | chr19 |
| 241 | DRG1 | 5.07E-128 | 0.37684024 | 0.507 | 0.144 | 1.49E-123 | chr22 |
| 12 | AFG3L2 | 5.39E-128 | 0.28294785 | 0.456 | 0.1 | 1.58E-123 | chr18 |
| 1 | A1BG | 6.87E-128 | 0.27927648 | 0.536 | 0.145 | 2.02E-123 | chr19 |
| 381 | KAZALD1 | 1.29E-127 | 0.28518294 | 0.429 | 0.083 | 3.78E-123 | chr10 |
| 668 | RSL24D1 | 2.81E-127 | 0.33606067 | 0.659 | 0.236 | 8.25E-123 | chr15 |
| 42 | ARL6IP5 | 3.17E-127 | 0.34540807 | 0.589 | 0.19 | 9.30E-123 | chr3 |
| 659 | RPA2 | 5.69E-127 | 0.29004085 | 0.519 | 0.141 | 1.67E-122 | chr1 |
| 377 | ITPA | 9.34E-127 | 0.26568901 | 0.561 | 0.168 | 2.74E-122 | chr20 |
| 566 | PKIG | 1.00E-126 | 0.25987392 | 0.498 | 0.124 | 2.94E-122 | chr20 |
| 486 | NBL1 | 1.53E-126 | 0.41586975 | 0.512 | 0.146 | 4.50E-122 | chr1 |
| 499 | NDUFB5 | 3.14E-126 | 0.30172387 | 0.58 | 0.187 | 9.21E-122 | chr3 |
| 158 | CHPF | 4.14E-126 | 0.29779571 | 0.504 | 0.132 | 1.21E-121 | chr2 |
| 861 | WDR45 | 4.37E-126 | 0.26799416 | 0.41 | 0.072 | 1.28E-121 | chrX |
| 730 | SLN | 9.71E-126 | 0.62088596 | 0.277 | 0.002 | 2.85E-121 | chr11 |
| 727 | SLC52A2 | 9.90E-126 | 0.30779699 | 0.555 | 0.168 | 2.90E-121 | chr8 |
| 641 | RCN3 | 1.87E-125 | 0.28103353 | 0.459 | 0.095 | 5.48E-121 | chr19 |
| 580 | POLE4 | 3.44E-125 | 0.33511642 | 0.598 | 0.211 | 1.01E-120 | chr2 |
| 661 | RPN1 | 3.62E-125 | 0.26757494 | 0.521 | 0.141 | 1.06E-120 | chr3 |
| 862 | WDR45B | 4.63E-125 | 0.33391254 | 0.617 | 0.211 | 1.36E-120 | chr17 |
| 459 | MRPS23 | 5.72E-125 | 0.32214524 | 0.583 | 0.195 | 1.68E-120 | chr17 |

| | | | | | | | |
|---|---|---|---|---|---|---|---|
| 331 | GSDMD | 6.93E-125 | 0.30370499 | 0.534 | 0.151 | 2.03E-120 | chr8 |
| 400 | LEPROT | 1.24E-124 | 0.36927782 | 0.549 | 0.175 | 3.65E-120 | chr1 |
| 542 | PDHA1 | 1.25E-124 | 0.51113853 | 0.55 | 0.192 | 3.65E-120 | chrX |
| 407 | LINC00632 | 1.26E-124 | 0.32007344 | 0.36 | 0.045 | 3.68E-120 | chrX |
| 496 | NDUFB1 | 1.26E-124 | 0.42705359 | 0.702 | 0.302 | 3.70E-120 | chr14 |
| 111 | C7orf50 | 2.03E-124 | 0.34330799 | 0.629 | 0.217 | 5.94E-120 | chr7 |
| 167 | CKLF | 2.19E-124 | 0.29131987 | 0.468 | 0.11 | 6.42E-120 | chr16 |
| 619 | PTK7 | 2.49E-124 | 0.30509733 | 0.497 | 0.132 | 7.29E-120 | chr6 |
| 560 | PHF5A | 2.77E-124 | 0.32426211 | 0.516 | 0.15 | 8.13E-120 | chr22 |
| 188 | COX19 | 3.61E-124 | 0.26371475 | 0.481 | 0.114 | 1.06E-119 | chr7 |
| 351 | HSF1 | 5.83E-124 | 0.31646577 | 0.625 | 0.221 | 1.71E-119 | chr8 |
| 89 | C14orf119 | 7.67E-124 | 0.26177199 | 0.52 | 0.143 | 2.25E-119 | chr14 |
| 332 | GSTK1 | 8.41E-124 | 0.26371272 | 0.478 | 0.113 | 2.47E-119 | chr7 |
| 348 | HMOX1 | 1.14E-123 | 0.3963545 | 0.338 | 0.034 | 3.34E-119 | chr22 |
| 631 | RAB3IL1 | 1.18E-123 | 0.25208352 | 0.368 | 0.052 | 3.46E-119 | chr11 |
| 368 | IGFBP6 | 1.52E-123 | 0.36189437 | 0.369 | 0.053 | 4.47E-119 | chr12 |
| 673 | S100A13 | 2.11E-123 | 0.56927999 | 0.728 | 0.416 | 6.19E-119 | chr1 |
| 380 | JTB | 3.23E-123 | 0.38723414 | 0.691 | 0.284 | 9.46E-119 | chr1 |
| 625 | PWP1 | 4.19E-123 | 0.26616112 | 0.508 | 0.132 | 1.23E-118 | chr12 |
| 123 | CCDC107 | 4.23E-123 | 0.27103208 | 0.501 | 0.127 | 1.24E-118 | chr9 |
| 357 | ICAM1 | 5.04E-123 | 0.28936549 | 0.285 | 0.008 | 1.48E-118 | chr19 |
| 763 | STARD7 | 5.81E-123 | 0.33772801 | 0.526 | 0.166 | 1.71E-118 | chr2 |
| 434 | MEAF6 | 7.41E-123 | 0.28874984 | 0.544 | 0.165 | 2.17E-118 | chr1 |
| 752 | SPPL3 | 8.26E-123 | 0.25294195 | 0.475 | 0.112 | 2.42E-118 | chr12 |
| 620 | PTN | 1.12E-122 | 1.01757238 | 0.784 | 0.441 | 3.27E-118 | chr7 |
| 7 | ACTR1B | 1.45E-122 | 0.29435018 | 0.482 | 0.124 | 4.26E-118 | chr2 |
| 726 | SLC44A2 | 1.57E-122 | 0.26677761 | 0.409 | 0.075 | 4.62E-118 | chr19 |
| 812 | TP53INP1 | 3.22E-122 | 0.26427927 | 0.314 | 0.022 | 9.44E-118 | chr8 |
| 398 | LAYN | 3.27E-122 | 0.31484328 | 0.396 | 0.064 | 9.61E-118 | chr11 |
| 628 | QSOX1 | 3.75E-122 | 0.26234979 | 0.414 | 0.076 | 1.10E-117 | chr1 |
| 672 | RUNX1T1 | 5.09E-122 | 0.43427001 | 0.57 | 0.182 | 1.49E-117 | chr8 |
| 312 | GALK1 | 5.45E-122 | 0.35906451 | 0.585 | 0.21 | 1.60E-117 | chr17 |
| 817 | TRIM55 | 6.33E-122 | 0.3840655 | 0.266 | 0 | 1.86E-117 | chr8 |
| 229 | DECR1 | 1.05E-121 | 0.42983308 | 0.619 | 0.248 | 3.09E-117 | chr8 |
| 221 | DCAF13 | 1.16E-121 | 0.41688846 | 0.666 | 0.286 | 3.41E-117 | chr8 |
| 259 | EIF3B | 2.37E-121 | 0.37335294 | 0.705 | 0.302 | 6.97E-117 | chr7 |
| 718 | SKP1 | 2.54E-121 | 0.51060017 | 0.806 | 0.486 | 7.45E-117 | chr5 |
| 860 | WDR34 | 3.70E-121 | 0.25327908 | 0.38 | 0.063 | 1.09E-116 | chr9 |
| 127 | CCL2 | 3.89E-121 | 0.56891003 | 0.356 | 0.047 | 1.14E-116 | chr17 |
| 37 | AQP1 | 4.03E-121 | 0.65008988 | 0.366 | 0.052 | 1.18E-116 | chr7 |

| | | | | | | | |
|---|---|---|---|---|---|---|---|
| 144 | CDCA4 | 7.63E-121 | 0.29683765 | 0.464 | 0.113 | 2.24E-116 | chr14 |
| 506 | NKAIN4 | 7.76E-121 | 0.32923857 | 0.348 | 0.043 | 2.28E-116 | chr20 |
| 809 | TNNI1 | 9.06E-121 | 0.55932782 | 0.281 | 0.009 | 2.66E-116 | chr1 |
| 72 | B3GALT6 | 1.54E-120 | 0.26451719 | 0.438 | 0.097 | 4.52E-116 | chr1 |
| 297 | FCGRT | 1.98E-120 | 0.37893309 | 0.613 | 0.218 | 5.80E-116 | chr19 |
| 21 | ALKBH7 | 2.29E-120 | 0.28699026 | 0.584 | 0.191 | 6.71E-116 | chr19 |
| 202 | CRNDE | 2.65E-120 | 0.32167075 | 0.532 | 0.159 | 7.78E-116 | chr16 |
| 820 | TROVE2 | 3.22E-120 | 0.29177709 | 0.518 | 0.149 | 9.45E-116 | chr1 |
| 636 | RARRES2 | 3.64E-120 | 0.53174037 | 0.64 | 0.253 | 1.07E-115 | chr7 |
| 248 | EBPL | 6.94E-120 | 0.31320648 | 0.524 | 0.153 | 2.04E-115 | chr13 |
| 73 | B4GALT2 | 8.27E-120 | 0.27588745 | 0.487 | 0.126 | 2.43E-115 | chr1 |
| 300 | FHL1 | 9.57E-120 | 0.26561592 | 0.448 | 0.1 | 2.81E-115 | chrX |
| 487 | NCALD | 1.13E-119 | 0.3656608 | 0.333 | 0.039 | 3.33E-115 | chr8 |
| 591 | PPDPF | 1.30E-119 | 0.47434451 | 0.746 | 0.347 | 3.81E-115 | chr20 |
| 691 | SEC61A1 | 1.37E-119 | 0.35216794 | 0.598 | 0.21 | 4.02E-115 | chr3 |
| 335 | HADHB | 1.63E-119 | 0.3255743 | 0.591 | 0.209 | 4.77E-115 | chr2 |
| 43 | ARMC1 | 1.63E-119 | 0.35407454 | 0.546 | 0.172 | 4.78E-115 | chr8 |
| 63 | ATP6AP2 | 1.64E-119 | 0.28676974 | 0.608 | 0.218 | 4.81E-115 | chrX |
| 378 | JAGN1 | 1.71E-119 | 0.3573451 | 0.525 | 0.166 | 5.00E-115 | chr3 |
| 714 | SIGIRR | 3.70E-119 | 0.32772507 | 0.487 | 0.136 | 1.08E-114 | chr11 |
| 11 | AEBP1 | 4.03E-119 | 0.62091479 | 0.705 | 0.299 | 1.18E-114 | chr7 |
| 568 | PLAC9 | 1.11E-118 | 0.62287426 | 0.407 | 0.094 | 3.27E-114 | chr10 |
| 799 | TMEM45A | 1.32E-118 | 0.29736002 | 0.399 | 0.075 | 3.88E-114 | chr3 |
| 35 | APMAP | 1.33E-118 | 0.26023938 | 0.472 | 0.12 | 3.91E-114 | chr20 |
| 651 | RNF181 | 1.70E-118 | 0.32258845 | 0.669 | 0.265 | 4.98E-114 | chr2 |
| 256 | EGLN3 | 1.70E-118 | 0.37208636 | 0.403 | 0.075 | 4.99E-114 | chr14 |
| 688 | SDHC | 3.29E-118 | 0.32698587 | 0.622 | 0.228 | 9.64E-114 | chr1 |
| 866 | XBP1 | 4.76E-118 | 0.39089945 | 0.637 | 0.242 | 1.40E-113 | chr22 |
| 590 | PPCS | 7.21E-118 | 0.26603082 | 0.498 | 0.135 | 2.12E-113 | chr1 |
| 87 | C12orf75 | 7.73E-118 | 0.28946587 | 0.441 | 0.101 | 2.27E-113 | chr12 |
| 533 | OXR1 | 9.08E-118 | 0.32488472 | 0.493 | 0.132 | 2.66E-113 | chr8 |
| 495 | NDUFA5 | 9.09E-118 | 0.27874628 | 0.637 | 0.223 | 2.67E-113 | chr7 |
| 396 | LAMTOR5 | 1.10E-117 | 0.46845856 | 0.711 | 0.365 | 3.23E-113 | chr1 |
| 227 | DDX21 | 1.24E-117 | 0.37286533 | 0.618 | 0.231 | 3.63E-113 | chr10 |
| 5 | ACTC1 | 1.53E-117 | 0.73105508 | 0.295 | 0.019 | 4.48E-113 | chr15 |
| 160 | CHRND | 2.28E-117 | 0.40695594 | 0.258 | 0 | 6.70E-113 | chr2 |
| 458 | MRPS18B | 2.90E-117 | 0.27605427 | 0.622 | 0.224 | 8.52E-113 | chr6 |
| 729 | SLIRP | 3.55E-117 | 0.40200214 | 0.722 | 0.343 | 1.04E-112 | chr14 |
| 19 | ALDH1A3 | 4.71E-117 | 0.37172784 | 0.433 | 0.098 | 1.38E-112 | chr15 |
| 543 | PDIA4 | 5.12E-117 | 0.3176121 | 0.623 | 0.228 | 1.50E-112 | chr7 |

| | | | | | | | |
|---|---|---|---|---|---|---|---|
| 310 | GADD45A | 6.11E-117 | 0.51346934 | 0.583 | 0.22 | 1.79E-112 | chr1 |
| 319 | GLO1 | 6.31E-117 | 0.31643719 | 0.633 | 0.237 | 1.85E-112 | chr6 |
| 317 | GIP | 6.66E-117 | 0.49029567 | 0.258 | 0.001 | 1.95E-112 | chr17 |
| 307 | FUOM | 1.03E-116 | 0.2667405 | 0.443 | 0.103 | 3.02E-112 | chr10 |
| 550 | PEPD | 1.32E-116 | 0.28091405 | 0.5 | 0.143 | 3.87E-112 | chr19 |
| 93 | C17orf58 | 1.91E-116 | 0.29461311 | 0.389 | 0.074 | 5.60E-112 | chr17 |
| 333 | GTF3A | 3.35E-116 | 0.36718431 | 0.644 | 0.258 | 9.83E-112 | chr13 |
| 128 | CCM2 | 6.59E-116 | 0.286897 | 0.56 | 0.182 | 1.93E-111 | chr7 |
| 45 | ASNA1 | 1.18E-115 | 0.25253487 | 0.515 | 0.155 | 3.48E-111 | chr19 |
| 341 | HK1 | 2.13E-115 | 0.25788065 | 0.463 | 0.114 | 6.25E-111 | chr10 |
| 146 | CDK16 | 3.10E-115 | 0.26613533 | 0.473 | 0.128 | 9.10E-111 | chrX |
| 232 | DGAT1 | 3.36E-115 | 0.32708096 | 0.525 | 0.155 | 9.86E-111 | chr8 |
| 650 | RNASEH2C | 6.82E-115 | 0.28608306 | 0.529 | 0.165 | 2.00E-110 | chr11 |
| 825 | TTYH3 | 1.39E-114 | 0.29996026 | 0.428 | 0.094 | 4.06E-110 | chr7 |
| 720 | SLC25A37 | 1.97E-114 | 0.45091497 | 0.477 | 0.127 | 5.79E-110 | chr8 |
| 800 | TMEM50A | 7.99E-114 | 0.33642867 | 0.64 | 0.241 | 2.35E-109 | chr1 |
| 592 | PPIC | 8.16E-114 | 0.29690556 | 0.525 | 0.164 | 2.39E-109 | chr5 |
| 32 | AP3D1 | 8.85E-114 | 0.27026358 | 0.554 | 0.179 | 2.60E-109 | chr19 |
| 404 | LIMA1 | 9.47E-114 | 0.28392685 | 0.482 | 0.13 | 2.78E-109 | chr12 |
| 611 | PSMB8 | 1.72E-113 | 0.37263448 | 0.571 | 0.203 | 5.03E-109 | chr6 |
| 824 | TSPO | 2.51E-113 | 0.48153527 | 0.747 | 0.405 | 7.35E-109 | chr22 |
| 666 | RRAGC | 6.31E-113 | 0.39494378 | 0.495 | 0.155 | 1.85E-108 | chr1 |
| 723 | SLC39A1 | 9.02E-113 | 0.31180729 | 0.608 | 0.231 | 2.65E-108 | chr1 |
| 34 | APH1A | 9.16E-113 | 0.33017187 | 0.65 | 0.253 | 2.69E-108 | chr1 |
| 142 | CDC25B | 9.42E-113 | 0.26243613 | 0.375 | 0.064 | 2.76E-108 | chr20 |
| 390 | KRT17 | 1.02E-112 | 1.02187924 | 0.318 | 0.038 | 2.98E-108 | chr17 |
| 515 | NPY | 4.52E-112 | 0.48513533 | 0.425 | 0.103 | 1.33E-107 | chr7 |
| 804 | TMEM98 | 4.57E-112 | 0.34271902 | 0.625 | 0.245 | 1.34E-107 | chr17 |
| 77 | BLCAP | 5.48E-112 | 0.2584717 | 0.492 | 0.137 | 1.61E-107 | chr20 |
| 521 | NUCB2 | 1.61E-111 | 0.34465801 | 0.551 | 0.188 | 4.72E-107 | chr11 |
| 402 | LGALS3BP | 2.05E-111 | 0.35971541 | 0.323 | 0.039 | 6.00E-107 | chr17 |
| 519 | NTRK2 | 2.21E-111 | 0.34924625 | 0.295 | 0.022 | 6.49E-107 | chr9 |
| 243 | DUSP1 | 2.58E-111 | 0.63553839 | 0.663 | 0.339 | 7.58E-107 | chr5 |
| 507 | NME3 | 3.59E-111 | 0.28578891 | 0.351 | 0.054 | 1.05E-106 | chr16 |
| 446 | MMADHC | 4.40E-111 | 0.25509047 | 0.592 | 0.206 | 1.29E-106 | chr2 |
| 593 | PPP1CB | 7.01E-111 | 0.32360113 | 0.62 | 0.239 | 2.06E-106 | chr2 |
| 608 | PSMA1 | 7.71E-111 | 0.31367778 | 0.707 | 0.328 | 2.26E-106 | chr11 |
| 847 | UXT | 9.45E-111 | 0.3235679 | 0.656 | 0.27 | 2.77E-106 | chrX |
| 124 | CCDC167 | 1.04E-110 | 0.31704599 | 0.583 | 0.213 | 3.06E-106 | chr6 |
| 86 | C11orf96 | 1.66E-110 | 0.62313436 | 0.407 | 0.102 | 4.87E-106 | chr11 |

| | | | | | | | |
|---|---|---|---|---|---|---|---|
| 38 | ARF3 | 1.89E-110 | 0.25124079 | 0.436 | 0.107 | 5.56E-106 | chr12 |
| 474 | MYADM | 2.17E-110 | 0.41454579 | 0.475 | 0.132 | 6.37E-106 | chr19 |
| 703 | SERPINI1 | 4.64E-110 | 0.34599199 | 0.355 | 0.061 | 1.36E-105 | chr3 |
| 412 | LOX | 6.66E-110 | 0.27857919 | 0.345 | 0.05 | 1.95E-105 | chr5 |
| 453 | MRPL33 | 8.71E-110 | 0.3030409 | 0.637 | 0.238 | 2.55E-105 | chr2 |
| 376 | ITGAE | 9.66E-110 | 0.26296731 | 0.533 | 0.167 | 2.83E-105 | chr17 |
| 881 | ZNF706 | 1.71E-109 | 0.47456604 | 0.762 | 0.453 | 5.02E-105 | chr8 |
| 387 | KLC1 | 1.74E-109 | 0.26338955 | 0.493 | 0.138 | 5.10E-105 | chr14 |
| 880 | ZNF593 | 2.12E-109 | 0.32971029 | 0.624 | 0.259 | 6.21E-105 | chr1 |
| 513 | NPTX2 | 2.47E-109 | 0.49435309 | 0.551 | 0.193 | 7.24E-105 | chr7 |
| 485 | NAGLU | 2.68E-109 | 0.3035461 | 0.41 | 0.094 | 7.87E-105 | chr17 |
| 526 | OAT | 4.18E-109 | 0.25419253 | 0.458 | 0.121 | 1.23E-104 | chr10 |
| 520 | NUCB1 | 7.61E-109 | 0.25040093 | 0.51 | 0.151 | 2.23E-104 | chr19 |
| 143 | CDC37 | 1.17E-108 | 0.33651508 | 0.691 | 0.308 | 3.43E-104 | chr19 |
| 260 | EIF3L | 1.64E-108 | 0.4311113 | 0.765 | 0.451 | 4.82E-104 | chr22 |
| 877 | ZNF22 | 1.97E-108 | 0.26934973 | 0.575 | 0.205 | 5.78E-104 | chr10 |
| 746 | SPATS2L | 2.44E-108 | 0.33606548 | 0.705 | 0.31 | 7.16E-104 | chr2 |
| 462 | MRPS7 | 3.20E-108 | 0.38531002 | 0.636 | 0.285 | 9.38E-104 | chr17 |
| 318 | GIPC1 | 3.47E-108 | 0.28447141 | 0.534 | 0.187 | 1.02E-103 | chr19 |
| 874 | ZFAND2A | 6.73E-108 | 0.39705897 | 0.597 | 0.229 | 1.97E-103 | chr7 |
| 360 | IER2 | 8.48E-108 | 0.5242587 | 0.767 | 0.391 | 2.49E-103 | chr19 |
| 293 | FBXO17 | 9.34E-108 | 0.25632958 | 0.491 | 0.143 | 2.74E-103 | chr19 |
| 731 | SMARCB1 | 1.05E-107 | 0.35137064 | 0.613 | 0.251 | 3.08E-103 | chr22 |
| 324 | GNPDA1 | 1.42E-107 | 0.31471061 | 0.451 | 0.125 | 4.16E-103 | chr5 |
| 129 | CCNB1IP1 | 1.62E-107 | 0.26821442 | 0.62 | 0.229 | 4.76E-103 | chr14 |
| 629 | RAB31 | 1.82E-107 | 0.30033319 | 0.424 | 0.104 | 5.34E-103 | chr18 |
| 122 | CBX6 | 2.37E-107 | 0.26771339 | 0.518 | 0.161 | 6.96E-103 | chr22 |
| 573 | PLIN3 | 2.56E-107 | 0.25752489 | 0.525 | 0.162 | 7.50E-103 | chr19 |
| 216 | CYC1 | 4.72E-107 | 0.4732586 | 0.771 | 0.515 | 1.39E-102 | chr8 |
| 379 | JPX | 4.86E-107 | 0.28616381 | 0.409 | 0.092 | 1.43E-102 | chrX |
| 383 | KDELR1 | 4.99E-107 | 0.39439405 | 0.755 | 0.382 | 1.46E-102 | chr19 |
| 698 | 9-Sep | 8.98E-107 | 0.34105592 | 0.577 | 0.218 | 2.63E-102 | chr17 |
| 240 | DPP7 | 9.31E-107 | 0.29371644 | 0.567 | 0.194 | 2.73E-102 | chr9 |
| 468 | MT1E | 9.40E-107 | 0.42132569 | 0.571 | 0.19 | 2.76E-102 | chr16 |
| 171 | CLU | 1.61E-106 | 0.36161339 | 0.521 | 0.167 | 4.72E-102 | chr8 |
| 238 | DNAJB11 | 2.34E-106 | 0.28978622 | 0.568 | 0.197 | 6.85E-102 | chr3 |
| 286 | FAM213A | 2.84E-106 | 1.33202173 | 0.493 | 0.205 | 8.34E-102 | chr10 |
| 195 | CPNE1 | 2.96E-106 | 0.25260345 | 0.623 | 0.232 | 8.69E-102 | chr20 |
| 6 | ACTN4 | 3.32E-106 | 0.29145105 | 0.6 | 0.232 | 9.74E-102 | chr19 |
| 366 | IGFBP4 | 4.09E-106 | 0.44145894 | 0.57 | 0.213 | 1.20E-101 | chr17 |

| | | | | | | | |
|---|---|---|---|---|---|---|---|
| 571 | PLEC | 5.25E-106 | 0.37172179 | 0.543 | 0.174 | 1.54E-101 | chr8 |
| 644 | RGS4 | 1.11E-105 | 0.5198206 | 0.301 | 0.031 | 3.25E-101 | chr1 |
| 584 | POLR2L | 2.61E-105 | 0.50282792 | 0.785 | 0.494 | 7.66E-101 | chr11 |
| 289 | FAM89B | 3.85E-105 | 0.2920956 | 0.585 | 0.216 | 1.13E-100 | chr11 |
| 676 | SAE1 | 5.14E-105 | 0.27578447 | 0.535 | 0.183 | 1.51E-100 | chr19 |
| 246 | DUT | 1.32E-104 | 0.25661561 | 0.597 | 0.212 | 3.88E-100 | chr15 |
| 577 | PLTP | 1.88E-104 | 0.26732894 | 0.393 | 0.089 | 5.52E-100 | chr20 |
| 490 | NDUFA1 | 2.86E-104 | 0.33657215 | 0.685 | 0.284 | 8.40E-100 | chrX |
| 423 | LYPLA2 | 4.89E-104 | 0.25071707 | 0.6 | 0.229 | 1.43E-99 | chr1 |
| 120 | CAST | 6.04E-104 | 0.27038126 | 0.521 | 0.163 | 1.77E-99 | chr5 |
| 546 | PDPN | 7.17E-104 | 0.31137133 | 0.352 | 0.067 | 2.10E-99 | chr1 |
| 80 | BOP1 | 1.70E-103 | 0.32177893 | 0.636 | 0.259 | 4.99E-99 | chr8 |
| 359 | IDS | 3.03E-103 | 0.25784295 | 0.351 | 0.061 | 8.88E-99 | chrX |
| 211 | CXCL3 | 3.27E-103 | 0.35206604 | 0.261 | 0.016 | 9.59E-99 | chr4 |
| 68 | ATXN10 | 3.79E-103 | 0.30621511 | 0.583 | 0.224 | 1.11E-98 | chr22 |
| 707 | SH3BP5 | 4.79E-103 | 0.33145325 | 0.465 | 0.144 | 1.40E-98 | chr3 |
| 589 | PPA1 | 9.79E-103 | 0.25970849 | 0.6 | 0.232 | 2.87E-98 | chr10 |
| 96 | C18orf32 | 1.06E-102 | 0.2690512 | 0.367 | 0.072 | 3.10E-98 | chr18 |
| 218 | CYTL1 | 1.10E-102 | 0.57827051 | 0.589 | 0.236 | 3.23E-98 | chr4 |
| 579 | PMAIP1 | 1.36E-102 | 0.35748208 | 0.556 | 0.192 | 3.98E-98 | chr18 |
| 632 | RAB42 | 1.44E-102 | 0.38712209 | 0.35 | 0.065 | 4.23E-98 | chr1 |
| 435 | MED10 | 2.53E-102 | 0.28514129 | 0.627 | 0.247 | 7.43E-98 | chr5 |
| 69 | AURKAIP1 | 2.89E-102 | 0.31918737 | 0.719 | 0.344 | 8.49E-98 | chr1 |
| 395 | LAMTOR1 | 3.09E-102 | 0.45202051 | 0.688 | 0.368 | 9.07E-98 | chr11 |
| 39 | ARHGDIA | 3.19E-102 | 0.33946724 | 0.683 | 0.328 | 9.37E-98 | chr17 |
| 234 | DGUOK | 3.78E-102 | 0.31250768 | 0.708 | 0.313 | 1.11E-97 | chr2 |
| 44 | ASAH1 | 4.57E-102 | 0.32620274 | 0.445 | 0.127 | 1.34E-97 | chr8 |
| 254 | EGFR | 5.01E-102 | 0.46300721 | 0.564 | 0.201 | 1.47E-97 | chr7 |
| 565 | PITX3 | 5.17E-102 | 0.33706096 | 0.254 | 0.014 | 1.52E-97 | chr10 |
| 725 | SLC40A1 | 1.70E-101 | 0.4091157 | 0.415 | 0.117 | 4.99E-97 | chr2 |
| 340 | HIST1H2BG | 1.71E-101 | 0.25352755 | 0.349 | 0.06 | 5.01E-97 | chr6 |
| 547 | PDXK | 2.00E-101 | 0.26918781 | 0.545 | 0.188 | 5.86E-97 | chr21 |
| 125 | CCDC34 | 8.61E-101 | 0.30935677 | 0.502 | 0.167 | 2.53E-96 | chr11 |
| 164 | CITED1 | 9.75E-101 | 0.26760119 | 0.275 | 0.025 | 2.86E-96 | chrX |
| 518 | NSMCE2 | 1.06E-100 | 0.25798201 | 0.542 | 0.184 | 3.10E-96 | chr8 |
| 854 | VKORC1 | 1.06E-100 | 0.31612762 | 0.632 | 0.261 | 3.12E-96 | chr16 |
| 193 | COX7B | 2.38E-100 | 0.39049728 | 0.72 | 0.359 | 6.99E-96 | chrX |
| 249 | ECH1 | 5.09E-100 | 0.29158586 | 0.635 | 0.269 | 1.49E-95 | chr19 |
| 177 | COL13A1 | 7.92E-100 | 0.26818545 | 0.359 | 0.069 | 2.32E-95 | chr10 |
| 769 | SURF4 | 1.12E-99 | 0.32850953 | 0.648 | 0.28 | 3.27E-95 | chr9 |

| | | | | | | | |
|---|---|---|---|---|---|---|---|
| 451 | MRC2 | 1.29E-99 | 0.36638201 | 0.561 | 0.203 | 3.79E-95 | chr17 |
| 311 | GADD45GIP1 | 2.27E-99 | 0.32390335 | 0.755 | 0.378 | 6.66E-95 | chr19 |
| 844 | USP11 | 2.85E-99 | 0.25001435 | 0.511 | 0.169 | 8.36E-95 | chrX |
| 516 | NR4A1 | 1.46E-98 | 0.2838621 | 0.569 | 0.222 | 4.27E-94 | chr12 |
| 172 | CMTM3 | 2.57E-98 | 0.25014255 | 0.525 | 0.175 | 7.54E-94 | chr16 |
| 414 | LOXL2 | 2.58E-98 | 0.4135302 | 0.496 | 0.163 | 7.58E-94 | chr8 |
| 823 | TSPAN5 | 7.81E-98 | 0.3065709 | 0.507 | 0.171 | 2.29E-93 | chr4 |
| 623 | PTRHD1 | 8.42E-98 | 0.30026437 | 0.633 | 0.278 | 2.47E-93 | chr2 |
| 749 | SPINK6 | 1.18E-97 | 1.35318327 | 0.313 | 0.056 | 3.45E-93 | chr5 |
| 819 | TRMT112 | 1.30E-97 | 0.35774882 | 0.788 | 0.432 | 3.81E-93 | chr11 |
| 583 | POLR2K | 1.43E-97 | 0.38814873 | 0.755 | 0.435 | 4.19E-93 | chr8 |
| 306 | FSCN1 | 3.55E-97 | 0.40558521 | 0.775 | 0.399 | 1.04E-92 | chr7 |
| 871 | ZC2HC1A | 5.18E-97 | 0.26132296 | 0.443 | 0.128 | 1.52E-92 | chr8 |
| 252 | EFR3A | 1.01E-96 | 0.25184513 | 0.413 | 0.105 | 2.96E-92 | chr8 |
| 337 | HEY1 | 1.17E-96 | 0.48613003 | 0.424 | 0.13 | 3.44E-92 | chr8 |
| 551 | PERP | 7.74E-96 | 0.36487868 | 0.526 | 0.191 | 2.27E-91 | chr6 |
| 81 | BRI3 | 1.24E-95 | 0.43358093 | 0.755 | 0.446 | 3.64E-91 | chr7 |
| 833 | TXNL4A | 1.48E-95 | 0.250918 | 0.674 | 0.301 | 4.34E-91 | chr18 |
| 832 | TXNIP | 1.53E-95 | 0.33175435 | 0.434 | 0.119 | 4.50E-91 | chr1 |
| 198 | CPVL | 2.30E-95 | 0.39451179 | 0.396 | 0.111 | 6.75E-91 | chr7 |
| 612 | PSMD13 | 2.47E-95 | 0.26352585 | 0.623 | 0.259 | 7.26E-91 | chr11 |
| 411 | LMNA | 4.09E-95 | 0.51368847 | 0.811 | 0.521 | 1.20E-90 | chr1 |
| 278 | EVA1B | 4.39E-95 | 0.31062862 | 0.648 | 0.3 | 1.29E-90 | chr1 |
| 17 | AKR1B1 | 5.48E-95 | 0.43277198 | 0.565 | 0.241 | 1.61E-90 | chr7 |
| 475 | MYC | 7.49E-95 | 0.26418671 | 0.485 | 0.151 | 2.20E-90 | chr8 |
| 782 | TMCO1 | 8.00E-95 | 0.25432352 | 0.666 | 0.27 | 2.35E-90 | chr1 |
| 401 | LGALS1 | 1.38E-94 | 0.32048228 | 0.888 | 0.639 | 4.04E-90 | chr22 |
| 724 | SLC3A2 | 2.25E-94 | 0.39827842 | 0.671 | 0.31 | 6.61E-90 | chr11 |
| 262 | EIF4EBP1 | 2.29E-94 | 0.33339591 | 0.648 | 0.305 | 6.71E-90 | chr8 |
| 554 | PGD | 2.82E-94 | 0.26333057 | 0.51 | 0.181 | 8.28E-90 | chr1 |
| 373 | INSIG1 | 2.92E-94 | 0.32135479 | 0.43 | 0.124 | 8.57E-90 | chr7 |
| 117 | CALU | 3.27E-94 | 0.41108136 | 0.739 | 0.382 | 9.61E-90 | chr7 |
| 669 | RTCB | 3.96E-94 | 0.25328045 | 0.552 | 0.215 | 1.16E-89 | chr22 |
| 478 | MYL6B | 4.88E-94 | 0.31091486 | 0.704 | 0.318 | 1.43E-89 | chr12 |
| 665 | RPUSD3 | 6.83E-94 | 0.30210737 | 0.442 | 0.137 | 2.00E-89 | chr3 |
| 635 | RAPSN | 8.80E-94 | 0.34807663 | 0.267 | 0.026 | 2.58E-89 | chr11 |
| 837 | UQCR10 | 3.53E-93 | 0.52974874 | 0.746 | 0.461 | 1.03E-88 | chr22 |
| 375 | ITGA10 | 6.61E-93 | 0.40617181 | 0.278 | 0.034 | 1.94E-88 | chr1 |
| 358 | ID1 | 6.76E-93 | 0.51319113 | 0.675 | 0.332 | 1.98E-88 | chr20 |
| 835 | UBE2T | 7.53E-93 | 0.28077429 | 0.399 | 0.109 | 2.21E-88 | chr1 |

| | | | | | | | |
|---|---|---|---|---|---|---|---|
| 347 | HMGN2 | 2.18E-92 | 0.60068381 | 0.822 | 0.574 | 6.39E-88 | chr1 |
| 711 | SHISA2 | 3.52E-92 | 0.43750749 | 0.388 | 0.113 | 1.03E-87 | chr13 |
| 639 | RBM38 | 5.20E-92 | 0.38545566 | 0.372 | 0.101 | 1.53E-87 | chr20 |
| 850 | VAT1 | 5.96E-92 | 0.29030881 | 0.42 | 0.126 | 1.75E-87 | chr17 |
| 483 | N4BP2L2 | 6.37E-92 | 0.36062331 | 0.654 | 0.263 | 1.87E-87 | chr13 |
| 849 | VASN | 8.45E-92 | 0.26494612 | 0.381 | 0.095 | 2.48E-87 | chr16 |
| 511 | NPDC1 | 9.26E-92 | 0.26472142 | 0.345 | 0.074 | 2.72E-87 | chr9 |
| 316 | GHITM | 6.16E-91 | 0.30765638 | 0.622 | 0.291 | 1.81E-86 | chr10 |
| 758 | SSR1 | 1.03E-90 | 0.2616198 | 0.599 | 0.241 | 3.02E-86 | chr6 |
| 609 | PSMA2 | 6.05E-90 | 0.29671456 | 0.764 | 0.425 | 1.77E-85 | chr7 |
| 537 | PABPC4 | 6.85E-90 | 0.27532004 | 0.652 | 0.279 | 2.01E-85 | chr1 |
| 827 | TUFM | 2.13E-89 | 0.26018688 | 0.697 | 0.312 | 6.24E-85 | chr16 |
| 572 | PLEKHA1 | 3.36E-89 | 0.26068733 | 0.42 | 0.116 | 9.86E-85 | chr10 |
| 199 | CRABP2 | 4.06E-89 | 0.44007515 | 0.713 | 0.349 | 1.19E-84 | chr1 |
| 722 | SLC2A3 | 4.22E-89 | 0.4774193 | 0.404 | 0.116 | 1.24E-84 | chr12 |
| 701 | SERPINF1 | 5.44E-89 | 0.33664432 | 0.641 | 0.305 | 1.60E-84 | chr17 |
| 786 | TMED4 | 1.24E-88 | 0.25977241 | 0.588 | 0.225 | 3.65E-84 | chr7 |
| 530 | OSR2 | 4.39E-88 | 0.30473969 | 0.419 | 0.124 | 1.29E-83 | chr8 |
| 613 | PSMD3 | 7.14E-88 | 0.27834353 | 0.622 | 0.272 | 2.10E-83 | chr17 |
| 646 | RHOB | 8.21E-88 | 0.37378569 | 0.657 | 0.326 | 2.41E-83 | chr2 |
| 791 | TMEM14B | 1.13E-87 | 0.26454752 | 0.616 | 0.245 | 3.32E-83 | chr6 |
| 811 | TP53I3 | 1.58E-87 | 0.28176196 | 0.465 | 0.165 | 4.65E-83 | chr2 |
| 344 | HLA-C | 2.24E-87 | 0.47901617 | 0.72 | 0.355 | 6.58E-83 | chr6 |
| 209 | CTSH | 2.63E-87 | 0.2624694 | 0.363 | 0.091 | 7.70E-83 | chr15 |
| 141 | CDC20 | 7.55E-87 | 0.346683 | 0.447 | 0.143 | 2.22E-82 | chr1 |
| 728 | SLC5A3 | 7.58E-87 | 0.86367505 | 0.518 | 0.194 | 2.22E-82 | chr21 |
| 205 | CTGF | 1.39E-86 | 0.50093255 | 0.586 | 0.277 | 4.09E-82 | chr6 |
| 161 | CIRBP | 1.60E-86 | 0.38873687 | 0.868 | 0.516 | 4.68E-82 | chr19 |
| 445 | MLLT11 | 2.95E-86 | 0.28877296 | 0.515 | 0.195 | 8.65E-82 | chr1 |
| 138 | CD81 | 3.85E-86 | 0.58838521 | 0.706 | 0.378 | 1.13E-81 | chr11 |
| 489 | NDRG1 | 4.07E-86 | 0.91826653 | 0.52 | 0.199 | 1.20E-81 | chr8 |
| 277 | ERP29 | 4.21E-86 | 0.30065163 | 0.699 | 0.374 | 1.24E-81 | chr12 |
| 33 | APEX1 | 6.21E-86 | 0.32036624 | 0.705 | 0.358 | 1.82E-81 | chr14 |
| 267 | EMC3 | 1.43E-85 | 0.25337436 | 0.506 | 0.184 | 4.18E-81 | chr3 |
| 220 | DBI | 5.20E-85 | 0.36175936 | 0.749 | 0.39 | 1.53E-80 | chr2 |
| 67 | ATRAID | 7.47E-85 | 0.33160652 | 0.728 | 0.391 | 2.19E-80 | chr2 |
| 634 | RALY | 1.06E-84 | 0.30691028 | 0.669 | 0.329 | 3.11E-80 | chr20 |
| 431 | MDFI | 2.72E-84 | 0.28464869 | 0.668 | 0.309 | 7.99E-80 | chr6 |
| 322 | GLUL | 4.19E-84 | 0.40170659 | 0.611 | 0.305 | 1.23E-79 | chr1 |
| 447 | MMP2 | 1.86E-83 | 0.32285649 | 0.591 | 0.255 | 5.47E-79 | chr16 |

| | | | | | | | |
|---|---|---|---|---|---|---|---|
| 739 | SNHG9 | 2.33E-83 | 0.2935173 | 0.339 | 0.08 | 6.82E-79 | chr16 |
| 22 | ANAPC11 | 2.81E-83 | 0.40087632 | 0.764 | 0.498 | 8.24E-79 | chr17 |
| 168 | CKS1B | 4.62E-82 | 0.30208198 | 0.667 | 0.32 | 1.35E-77 | chr1 |
| 391 | KRT8 | 8.73E-82 | 0.34781566 | 0.274 | 0.044 | 2.56E-77 | chr12 |
| 433 | MDM4 | 1.31E-81 | 0.27141155 | 0.393 | 0.11 | 3.83E-77 | chr1 |
| 558 | PGM2L1 | 1.56E-81 | 0.27793459 | 0.446 | 0.149 | 4.58E-77 | chr11 |
| 223 | DDIT3 | 4.56E-81 | 0.36742263 | 0.594 | 0.277 | 1.34E-76 | chr12 |
| 498 | NDUFB4 | 1.24E-80 | 0.25291088 | 0.703 | 0.333 | 3.63E-76 | chr3 |
| 196 | CPNE3 | 1.31E-80 | 0.2652684 | 0.62 | 0.287 | 3.83E-76 | chr8 |
| 314 | GDF15 | 1.38E-80 | 0.81638424 | 0.254 | 0.033 | 4.04E-76 | chr19 |
| 538 | PARK7 | 1.64E-80 | 0.30614505 | 0.796 | 0.511 | 4.82E-76 | chr1 |
| 834 | UBE2C | 1.67E-80 | 0.34078205 | 0.464 | 0.162 | 4.90E-76 | chr20 |
| 145 | CDK1 | 2.75E-80 | 0.28850331 | 0.351 | 0.091 | 8.06E-76 | chr10 |
| 253 | EGFL7 | 2.78E-80 | 0.26502812 | 0.517 | 0.193 | 8.15E-76 | chr9 |
| 765 | STOML2 | 3.34E-80 | 0.25481042 | 0.662 | 0.329 | 9.81E-76 | chr9 |
| 744 | SPARCL1 | 4.28E-80 | 0.80379769 | 0.347 | 0.091 | 1.26E-75 | chr4 |
| 602 | PRKDC | 6.85E-80 | 0.2526254 | 0.625 | 0.268 | 2.01E-75 | chr8 |
| 236 | DLC1 | 7.96E-80 | 0.28295638 | 0.391 | 0.115 | 2.33E-75 | chr8 |
| 178 | COL4A2 | 2.61E-79 | 0.47067666 | 0.612 | 0.267 | 7.66E-75 | chr13 |
| 500 | NDUFB8 | 3.04E-78 | 0.3216737 | 0.731 | 0.433 | 8.91E-74 | chr10 |
| 753 | SQLE | 5.51E-78 | 0.33557043 | 0.611 | 0.267 | 1.62E-73 | chr8 |
| 320 | GLRX5 | 1.53E-77 | 0.25231773 | 0.63 | 0.298 | 4.48E-73 | chr14 |
| 783 | TMED10 | 1.37E-76 | 0.25911295 | 0.727 | 0.402 | 4.01E-72 | chr14 |
| 269 | EMP2 | 4.74E-76 | 0.26776297 | 0.541 | 0.213 | 1.39E-71 | chr16 |
| 191 | COX7A2 | 1.09E-75 | 0.31068257 | 0.806 | 0.478 | 3.19E-71 | chr6 |
| 382 | KCNMA1 | 1.38E-75 | 0.29463406 | 0.26 | 0.041 | 4.06E-71 | chr10 |
| 76 | BIRC5 | 3.03E-75 | 0.32979872 | 0.458 | 0.17 | 8.90E-71 | chr17 |
| 690 | SEC13 | 3.95E-75 | 0.34671875 | 0.609 | 0.316 | 1.16E-70 | chr3 |
| 26 | ANKRD37 | 1.00E-74 | 0.29808221 | 0.349 | 0.093 | 2.94E-70 | chr4 |
| 308 | FXYD5 | 3.11E-74 | 0.37836351 | 0.452 | 0.191 | 9.11E-70 | chr19 |
| 428 | MAP1LC3B | 3.52E-74 | 0.25468348 | 0.644 | 0.289 | 1.03E-69 | chr16 |
| 31 | AP2M1 | 4.61E-74 | 0.35064139 | 0.709 | 0.44 | 1.35E-69 | chr3 |
| 273 | ENY2 | 6.71E-74 | 0.42581353 | 0.831 | 0.627 | 1.97E-69 | chr8 |
| 606 | PRSS35 | 9.66E-74 | 0.28965109 | 0.305 | 0.072 | 2.83E-69 | chr6 |
| 642 | RGS16 | 1.92E-73 | 0.29160311 | 0.483 | 0.187 | 5.63E-69 | chr1 |
| 581 | POLR2G | 4.71E-73 | 0.25885661 | 0.649 | 0.341 | 1.38E-68 | chr11 |
| 425 | MAGED1 | 1.13E-72 | 0.28347046 | 0.683 | 0.372 | 3.32E-68 | chrX |
| 870 | YWHAZ | 1.15E-72 | 0.38499981 | 0.832 | 0.619 | 3.38E-68 | chr8 |
| 14 | AHI1 | 1.44E-72 | 0.25459424 | 0.457 | 0.164 | 4.22E-68 | chr6 |
| 878 | ZNF395 | 6.53E-71 | 0.34410065 | 0.291 | 0.067 | 1.92E-66 | chr8 |

| | | | | | | | |
|---|---|---|---|---|---|---|---|
| 808 | TNMD | 7.16E-70 | 0.79846137 | 0.318 | 0.09 | 2.10E-65 | chrX |
| 539 | PARP1 | 1.43E-69 | 0.41491732 | 0.571 | 0.29 | 4.19E-65 | chr1 |
| 760 | SSR3 | 1.69E-69 | 0.26943238 | 0.671 | 0.329 | 4.95E-65 | chr3 |
| 559 | PHF14 | 1.69E-69 | 0.2512346 | 0.718 | 0.363 | 4.96E-65 | chr7 |
| 681 | SCD | 4.90E-69 | 0.27231687 | 0.408 | 0.143 | 1.44E-64 | chr10 |
| 346 | HM13 | 5.92E-69 | 0.25685368 | 0.663 | 0.328 | 1.74E-64 | chr20 |
| 242 | DSTN | 2.77E-68 | 0.27277176 | 0.724 | 0.372 | 8.11E-64 | chr20 |
| 261 | EIF4A3 | 9.79E-68 | 0.27236874 | 0.605 | 0.304 | 2.87E-63 | chr17 |
| 575 | PLOD2 | 2.47E-67 | 0.267225 | 0.479 | 0.189 | 7.24E-63 | chr3 |
| 27 | ANXA1 | 1.42E-66 | 0.41234665 | 0.556 | 0.255 | 4.15E-62 | chr9 |
| 671 | RTN4 | 2.12E-66 | 0.37519553 | 0.77 | 0.48 | 6.22E-62 | chr2 |
| 294 | FBXO32 | 3.34E-66 | 0.25576532 | 0.266 | 0.057 | 9.79E-62 | chr8 |
| 541 | PDGFA | 2.00E-65 | 0.27410205 | 0.265 | 0.057 | 5.86E-61 | chr7 |
| 292 | FBN2 | 8.51E-64 | 0.3519841 | 0.419 | 0.163 | 2.50E-59 | chr5 |
| 610 | PSMB7 | 5.08E-63 | 0.25054858 | 0.714 | 0.478 | 1.49E-58 | chr9 |
| 116 | CALD1 | 7.59E-63 | 0.39232277 | 0.843 | 0.543 | 2.23E-58 | chr7 |
| 352 | HSPA1A | 8.48E-63 | 0.33750138 | 0.64 | 0.395 | 2.49E-58 | chr6 |
| 633 | RAD21 | 1.19E-62 | 0.29240767 | 0.762 | 0.448 | 3.49E-58 | chr8 |
| 637 | RBFOX2 | 1.32E-62 | 0.29482511 | 0.672 | 0.366 | 3.87E-58 | chr22 |
| 417 | LTBP1 | 1.57E-62 | 0.33271932 | 0.499 | 0.216 | 4.61E-58 | chr2 |
| 742 | SOCS3 | 2.38E-62 | 0.28670748 | 0.409 | 0.153 | 6.98E-58 | chr17 |
| 245 | DUSP6 | 1.38E-61 | 0.36856359 | 0.597 | 0.321 | 4.06E-57 | chr12 |
| 527 | OGN | 1.98E-60 | 0.51425918 | 0.335 | 0.113 | 5.81E-56 | chr9 |
| 776 | TCF25 | 2.90E-60 | 0.2570368 | 0.722 | 0.394 | 8.50E-56 | chr16 |
| 104 | C1R | 3.99E-60 | 0.25644458 | 0.348 | 0.12 | 1.17E-55 | chr12 |
| 175 | COCH | 7.86E-60 | 0.26505675 | 0.32 | 0.103 | 2.31E-55 | chr14 |
| 761 | ST13 | 2.62E-59 | 0.2801003 | 0.771 | 0.5 | 7.69E-55 | chr22 |
| 813 | TPI1 | 3.25E-59 | 0.3434244 | 0.873 | 0.626 | 9.52E-55 | chr12 |
| 585 | POMP | 3.40E-59 | 0.28156171 | 0.796 | 0.587 | 9.99E-55 | chr13 |
| 869 | YBX3 | 3.25E-58 | 0.30737028 | 0.769 | 0.492 | 9.54E-54 | chr12 |
| 276 | ERH | 3.98E-58 | 0.26170829 | 0.774 | 0.525 | 1.17E-53 | chr14 |
| 268 | EMILIN1 | 5.37E-58 | 0.25636477 | 0.625 | 0.307 | 1.58E-53 | chr2 |
| 166 | CKB | 6.27E-58 | 0.46625907 | 0.674 | 0.407 | 1.84E-53 | chr14 |
| 864 | WIF1 | 9.70E-58 | 0.28724417 | 0.339 | 0.118 | 2.85E-53 | chr12 |
| 272 | ENPP2 | 3.88E-55 | 0.40501273 | 0.447 | 0.201 | 1.14E-50 | chr8 |
| 467 | MT-ND6 | 5.89E-54 | 0.27084886 | 0.613 | 0.298 | 1.73E-49 | chrM |
| 600 | PRDX6 | 6.63E-54 | 0.36805847 | 0.725 | 0.471 | 1.94E-49 | chr1 |
| 389 | KPNA2 | 1.21E-53 | 0.41428464 | 0.575 | 0.312 | 3.55E-49 | chr17 |
| 271 | ENO2 | 1.33E-52 | 0.26306416 | 0.323 | 0.114 | 3.90E-48 | chr12 |
| 734 | SMS | 4.87E-52 | 0.25150981 | 0.521 | 0.268 | 1.43E-47 | chrX |

| | | | | | | | |
|---|---|---|---|---|---|---|---|
| 364 | IFITM3 | 1.83E-50 | 0.28580437 | 0.784 | 0.54 | 5.38E-46 | chr11 |
| 674 | S100A16 | 8.75E-50 | 0.36530663 | 0.38 | 0.171 | 2.57E-45 | chr1 |
| 664 | RPS26 | 1.88E-49 | 0.28822766 | 0.868 | 0.656 | 5.52E-45 | chr12 |
| 176 | COL11A1 | 5.73E-49 | 0.31141224 | 0.521 | 0.262 | 1.68E-44 | chr1 |
| 517 | NRN1 | 1.26E-39 | 0.3225009 | 0.409 | 0.199 | 3.70E-35 | chr6 |
| 464 | MT-ND2 | 1.62E-37 | 0.56552153 | 0.933 | 0.727 | 4.76E-33 | chrM |
| 865 | WSB1 | 1.05E-36 | 0.29302085 | 0.739 | 0.415 | 3.09E-32 | chr17 |
| 852 | VEGFA | 1.18E-36 | 0.77711692 | 0.456 | 0.249 | 3.47E-32 | chr6 |
| 514 | NPW | 1.11E-34 | 0.25087623 | 0.726 | 0.422 | 3.25E-30 | chr16 |
| 821 | TRPS1 | 2.33E-24 | 0.25820324 | 0.613 | 0.359 | 6.85E-20 | chr8 |
| 466 | MT-ND5 | 2.28E-21 | 0.44677319 | 0.903 | 0.649 | 6.68E-17 | chrM |
| 367 | IGFBP5 | 5.12E-17 | 0.27529091 | 0.724 | 0.453 | 1.50E-12 | chr2 |
| 465 | MT-ND4L | 1.07E-10 | 0.33910135 | 0.755 | 0.499 | 3.14E-06 | chrM |

**Supplemental Table 5: Marker genes of cells that demonstrate gain of Chr8q.**

### Materials & Methods

*Human subjects and patient-derived xenograft (PDX) tumor models*

Human tumor samples were collected under IRB-approved protocols (#201203042 at Washington University and J1649 at Johns Hopkins University), and following informed patient consent. A total of 2-4 pieces of tumor tissue were collected from each surgical specimen, immediately following resection, while the tumor was being processed in pathology. Tissues were placed into DMEM medium during transport to the laboratory, and then used for implantation into mice, histology, and sequencing of the parental tumor. Tissue for histology was fixed in 10% formalin, and stained with hematoxylin and eosin (H&E) to appropriately characterize the morphology of the implanted tumor. Tumor tissue was implanted subcutaneously into the flank of five to six-week old NSG mice (Jackson Lab). The mean elapsed time for engraftment varied from 21 to 90 days and engraftment occurred in 2-4 mice. Engraftment success of the PDX tumor model was defined by the ability for the line to be serially transplanted. Passage 5 was used for comparison studies.

*Histological evaluation of primary and PDX tumors*

Sections (6 $\mu$m) were prepared from formalin-fixed paraffin-embedded blocks for immunohistochemical stains (IHC) with adequate controls. IHC was performed using the Avidin/Biotin blocking kit (Vector SP-2001) staining with antibodies against S100, Ki-67, and Col4A as previously described(1).

*Whole Exome Sequencing (WES), Whole Genome Sequencing (WGS), RNA Sequencing (RNA-Seq) Library Construction and Sequencing*

Each tumor and xenograft had two enriched libraries constructed (n = 20), and the normal germline samples had a single enriched library constructed (n = 10). Exome libraries were captured with an IDT exome reagent, then pooled with a WGS library for sequencing on an Illumina HiSeq4000 with at least 1000x coverage. RNA was prepared with a TrueSeq stranded total RNA library kit, then sequenced on an Illumina HISeq4000 at an average of 52M reads/sample. All WES/WGS/RNA-Seq samples were trimmed and quality controlled via Trim Galore (https://www.bioinformatics.babraham.ac.uk/projects/trim_galore/).

*Whole Exome Sequencing Data Analysis*

WES and WGS data were aligned against reference sequence hg38 via BWA-MEM (2) with Base Quality Score Recalibration (BQSR). For PDX sequence data, Disambiguate v1.0(3) was used to filter out mouse-derived reads using mouse (GRCm38.p6, GENCODE release M19) and human (hg38) reference genomes, and the resulting reads were deduplicated. Structural variants (SVs) and large indels were detected using Manta (4). SNVs and small indels were detected using VarScan2 (5), Strelka2 (6), MuTect2 (7), and Pindel (8) via the somatic pipelines available at https://github.com/genome/analysis-workflows, which includes best-practices variant filtering and annotation with VEP (Variant Effect Predictor, version 95) (9). Manual review was used to remove additional sequencing artifacts. Germline and somatic variants reported by the variant-detection pipeline were compared, and any intersecting variants were removed from the somatic variant gene list, thus filtering out the germline variants. Common variants found in the 1000 Genomes MAF (minor allele frequency) > 0.05 were filtered out. Waterfall somatic variant plots were created with GenVisR (10) by including somatic variants that occurred in each area.

*Inference of copy number variations on Whole Genome Sequencing Data*

CNVkit was used to infer and visualize copy number from high-throughput WGS sequencing data. Coverage for each bait position in the exome reagent was calculated, then segments of constant copy number were identified using circular binary segmentation. Data were plotted to provide visualization of CNVs. John Hopkin's CNV data from PN was directly imported from Synapse.

*Fluorescent in situ hybridization*

Interphase FISH (fluorescence in situ hybridization) was performed on formalin fixed paraffin-embedded tissue sections cut at a thickness of 5-μm on positively charged microscope slides. The paraffin was removed from the sections with three washes of 5 minutes each in CitriSolve. The slides were then hydrated in two washes of absolute ethanol for 1 minute each and allowed to air dry. The slides were processed through a pretreatment solution of sodium thiocyanate which had been pre-heated to 80°C. After a 3-minute wash in distilled water, the tissue was digested in protease solution (pepsin in 0.2N HCl) for 15 minutes at 37°C, followed by another 3-minute wash in distilled water. The slides were allowed to air dry after which they were dehydrated by passing through consecutive 70%, 85%, and 100% ethanol solutions for 1 minute each. The slides were again allowed to air dry before applying prepared probe mixture. Probes used were a centromeric-enumerating probe - Vysis CEP 8 (D8Z2) SpectrumGreen combined with locus-specific probe - Vysis LSI MYC SpectrumOrange (Abbott Molecular, Des Plaines, IL). Probes were diluted at a concentration of 1:50 in *t*DenHyb-2 hybridization buffer (Insitus Biotechnologies, Inc., Albuquerque, NM) and well-mixed. Next, probe in buffer was applied to the appropriate slide to cover the tissue section and the section was cover-slipped. Co-denaturation was achieved by incubating the slides at 73°C for 5 minutes in a slide moat. Hybridization occurred by transferring the slides to a 37°C slide light-shielded, humid slide moat overnight. Post-hybridization, the coverslips were removed and the slides immersed in 75°C wash solution (2XSSC/0.3%NP40) for 2 minutes followed by a 1-minute wash in a jar containing the same solution at room temperature. The slides were allowed to air dry in the dark and were then counterstained with 10μl of DAPI II (Abbott Molecular, Inc.). Slides were examined using an Olympus BX61 fluorescent microscope with appropriate filters for SpectrumOrange, SpectrumGreen, and the DAPI counterstain. The signal patterns were documented using a JAI Progressive Scan camera and CytoVision Imaging System.

*Single-cell RNAseq*

Single-cell RNA sequencing was performed on the Chromium controller (10X Genomics) and sequenced using Illumina NextSeq sequencer. MPNST PDXs were minced in a cell culture dish and immediately dissociated in tumor dissociation media (DMEM supplemented with 10% FBS, 100 U/mL penicillin streptomycin solution, dispase II, collagenase I and DNase I) on a gentleMACS dissociator (Miltenyi Biotec). Single cell suspension was counted using both Cellometer Auto 2000 fluorescent viability cell counter (Nexcelom) and Luna-FL dual fluorescent cell counter (Logos Biosystems). The manufacturer's protocol was followed with a target capture of 6000 live cells from each sample. WU-386, WU-225, WU-368, JH2-023, JH2-002, WU-356 and WU-436 were processed using the 3' Library and Gel Bead Kit v2 (10X Genomics). PDX JH2-031 was processed using the 3' Library and Gel Bead Kit v3 (10X Genomics). Each sample was processed on an independent Chromium Single Cell A Chip. Cell partitioning completed successfully with uniform emulsion consistency and was followed with reverse transcription in a thermal cycler. All subsequent steps of library preparation and quality control were performed as described in the 10X Genomics 3' Single cell User Guide. Gene expression libraries were sequenced using the NextSeq High Output 150-cycle v2.5 Flow Cell.

*Single-cell RNAseq data analysis*

The standard 10X Genomics Cell Ranger (version 3.1.0) pipeline was used to extract FASTQ files and to perform data processing. Sequenced reads were aligned to the 10X Genomics provided human and mouse reference (refdata-cellranger-GRCh38-and- mm10-3.1.0). Human genome aligned reads were used in the downstream analysis. Each sample was initially processed separately and then integrated. For individual tumor analysis, "filtered_feature_bc_matrix" was imported using the Read10X function in Seurat (version 3.0) and a Seurat object was created keeping all genes expressed in more than 4 cells and all cells with more than 200 genes detected. Cell were further filtered after an initial quality control step in Seurat to retain cells with unique gene counts that were between 200 to 6000 and the percentage of mitochondrial genes that was lower than 25%. After removing the unwanted cells, the data was normalized using the "LogNormalize" method with a scale factor of 10000 and scaled to remove the unwanted sources of variation from mitochondrial gene content and number of detected unique molecules in each cell. The top 2000 variable genes were calculated using the FindVariableFeatures function in Seurat and used in the following principle component analysis (PCA) to reduce dimensionality of the data. The first forty PCs were used to find clusters in the data with the resolution set to 1.2. The resulting clusters were visualized in a two-dimensional UMAP representation with distinct clusters annotated. All 10X Genomics Cell Ranger processed, human hg38 genome annotated count matrices (one from each PDX) were imported into Seurat and integrated into one dataset following the standard integration pipeline. The top 2000 variably expressed genes of each PDX were used in the anchor-based integration with the first 30 PCs. Unsupervised clustering was performed and the 2-dimensional UMAP representation was used to exhibit the cell types detected. The FindAllMarkers function was used to identify conserved cell markers when comparing one cell cluster to the other clusters all together. Seurat (version 3.0) was used for the analysis in R (version 3.6.0) and data visualization.

*Inference of copy number variations and clonality analysis*

Copy number variations of each PDX were inferred from single-cell RNAseq using the R package inferCNV (https://github.com/broadinstitute/infercnv). Raw counts of each PDX were extracted from the Seurat object of individual samples following the recommendations in the "Using 10x data" section of inferCNV. A single-cell RNAseq dataset of plexiform neurofibroma that is composed of 19,000 cells randomly selected from 4 human primary plexiform neurofibromas was used as normal reference cells. The following parameters were used for the inferCNV analysis: cutoff=0.1 to use the genes with a mean number of counts larger than 0.1, denoise=T to obtain the residual CNV signal by subtracting the mean CNV signal from the normal cells to, and HMM=T to apply the default Hidden Markov Model. To rule out false-positive CNVs detected by the Hidden Markov Model, a Bayesian latent mixture model is implemented to identify the posterior probabilities of alteration status in each cell with each gene or CNA regions. The default probability threshold of 0.5 was used. To determine the clonality within each tumor due to the intratumoral heterogeneity, the "subcluster" analysis mode was used with the tumor_subcluster_partition_method set to "random_trees" and tumor_subcluster_pval set to 0.05. A six-state CNV model was adapted by inferCNV and each detected CNV was predicted to be in one of the following CNV levels: State 1, complete loss; State 2, loss of one copy; State 3, neutral; State 4, addition of one copy; State 5, addition of two copies; State 6 addition of more than 2 copies. The CNV was considered as "loss" for States 1 and 2 or "gain" for States 4, 5 and 6 events. Each CNV events detected was summarized in large-scale genomic regions and attributed to the chromosome arm levels according to the hg38 cytoband information (http://hgdownload.cse.ucsc.edu/goldenpath/hg38/database/). The identified CNV types and calculated percentage of cells within each CNV type were plotted as a phylogenetic tree. The Uphyloplot2 (https://github.com/harbourlab/UPhyloplot2) was used to visualize the clonality determined by inferCNV analysis of each PDX. CNV data was processed in R (version 3.6.2) and the summary of each tumor is presented in **Supplemental Table 4**. All detected CNV events were summarized and the percentage of cells exhibiting each characteristic CNV event in each PDX was plotted in a heatmap using R package (ComplexHeatmap).

*Data storage and Sharing*

All shared data are stored in the SYNAPSE repository: doi:10.7303/syn11638893 and will be made available to all researchers.
